# Elimination Capacity Determines Outcome in Amatoxin Mushroom Poisoning-Induced Acute Liver Failure: A Globally Applicable Management Framework

**DOI:** 10.64898/2026.03.05.26345777

**Authors:** S Todd Mitchell, Daniel A Spyker, Glenn Robbins, Barry H Rumack

## Abstract

Amatoxin mushroom poisoning causes acute liver failure; progression from cholera-like hypovolemia resolution to fatal multisystem collapse occurs unpredictably. We conducted a prospective single-arm multicenter trial (N=99) investigating proactive toxicokinetic-based management: sustained hydration for renal protection and clearance preservation, fasting plus octreotide to arrest enterohepatic circulation, and intravenous silibinin to block hepatic reuptake. Transplant-free recovery: 98.8% among protocol-adherent patients (n=86). Multivariable analysis: sustained hydration (P<0.001) and silibinin (P=0.003) predicted survival; octreotide reduced coagulopathy recovery time (P=0.01); N-acetylcysteine and charcoal achieved neither. Conventional MELD score–outcome relationships vanished in protocol-adherent patients, suggesting management modified the natural history. Poor outcomes followed a reproducible sequence—hydration withdrawal, lactate rebound, oliguric nephropathy, multisystem failure—kidney failure, not liver, precipitated collapse. These findings establish renal clearance preservation and gallbladder sequestration as central outcome determinants, supporting a globally applicable management framework substituting gallbladder drainage when silibinin is unavailable or likely to fail.

ClinicalTrials.gov identifier: NCT00915681.

## INTRODUCTION

Amatoxin mushroom poisoning presents a distinctive clinical paradox: hemodynamic and gastrointestinal symptoms recede with post-presentation volume replacement, yet some patients precipitously decline into multisystem failure hours to days later.^1,2^ This unexpected transition from symptomatic improvement to fatal collapse suggests mechanisms beyond primary hepatic injury alone. While outcome risk correlates with ingested dose,^1–3^ abrupt deterioration can follow even small ingestions; dose alone does not reliably predict clinical trajectory.^4–6^ This unpredictability contributes to the considerable mortality still documented in tertiary transplant centers as well as the resource-limited settings where most poisoning fatalities occur.^7–10^ The determinants of recovery or collapse remain incompletely defined; the prevailing assumption that hepatic injury drives kidney failure may have the causal relationship inverted. The association between toxin elimination capacity and progression to severe ALF manifestations has not been systematically evaluated.

Amatoxin inhibits RNA polymerase II, causing transcriptional arrest and apoptosis in enterocytes, hepatocytes, and tubular epithelium.^2^ Thermostable, water-soluble, metabolically inert, and eliminated intact, its toxicokinetics feature rapid absorption, minimal protein binding, low distribution volume, and creatinine-like clearance with >80% of the absorbed dose recoverable in urine within hours of ingestion.^11–15^ Organic anion transporter polypeptide 1B3 (OATP1B3) actively mediates sinusoidal hepatic uptake,^16^ making the liver the target organ and ALF the cause of death. The hepatocellular fraction not bound to RNA polymerase II is quickly excreted into bile,^11–15,17^ which accumulates in the gallbladder.^18^ Meal-triggered gallbladder contraction releases this concentrated bile into the intestine,^19^ enabling renewed amatoxin absorption, hepatic re-uptake, and hepatotoxic exacerbation. By contrast, nephrotoxicity is concentration– and time-dependent, developing from tubular stasis during the prolonged prerenal interval between repetitive vomiting and cholera-like diarrhea-induced hypovolemia onset and post-presentation volume restoration.^12,20–22^ Absorption is species-dependent and^,11–13,23,24^ like hepatic uptake, may be transporter-facilitated. Rodents absorb amatoxin poorly,^2,23,24^ an underappreciated difference with higher mammals that confounds interpretation of murine studies seeking antidotal validation.^25^ Poisonings unfold in four phases: 1) presymptomatic latency (6–24 hours); 2) dehydrating gastrointestinal; 3) postsymptomatic pseudo-recovery; and 4) fulminant multisystem failure. The gastrointestinal syndrome is caused by amatoxin itself, not phallotoxins; parenteral α-amanitin reproduces it in dogs.^23,24^

Silibinin dihemisuccinate (Legalon SIL, Viatris), in routine clinical use across Europe, is a highly water-soluble intravenous preparation derived from the common milk thistle (*Silybum marianum*) that inhibits sinusoidal OATP1B3-mediated hepatic uptake,^16,26^ diverting amatoxin away from hepatocellular uptake and into the hepatic venous outflow for glomerular filtration and urinary elimination.^26^ Beagle studies and retrospective data led to European approvals ∼40 years ago,^27,28^ but prospective evaluation never followed,^26^ and treatment failures remain unexplained.^7–10^ Oral silymarin preparations have poor bioavailability, limiting systemic delivery, and are not regulated; a recent study failed to establish efficacy.^29^

We hypothesized that impaired elimination capacity, rather than inexorable primary injury, is the principal determinant of clinical outcome—reflecting a kinetic balance between renal clearance and hepatic uptake. Under this framework, the gallbladder serves as the critical toxin accumulation reservoir: when renal clearance is preserved, most enterohepatic circulation (EHC)-reintroduced systemic amatoxin (>80%) undergoes urinary elimination;^11–15,21,22,24^ when clearance fails, this reintroduced systemic fraction distributes to the liver instead,^30^ precipitating the collapse characteristic of fatal poisoning. We further hypothesized that proactive management designed to preserve renal clearance and suppress EHC—pending silibinin infusion—would improve outcomes by maintaining the elimination pathways upon which antidotal efficacy depends.^31^ Because randomized controlled trials for rapidly fatal poisoning antidotes are neither ethically nor practically feasible,^32,33^ and building upon the unexpectedly favorable clinical outcomes in severe cases treated under Emergency Investigational New Drug (IND) authorization,^34^ we undertook a prospective multicenter single-arm study to evaluate the relationship between toxin elimination capacity and transplant-free recovery in amatoxin-induced acute liver failure.

## METHODS

### Human Subjects Protection

The study protocol and amendments were approved by the Catholic Healthcare West Institutional Review Board (Santa Cruz, CA); participating hospitals obtained local IRB authorization. All participants (or parents/legal guardians for minors) provided written informed consent.

### Study Design

This was a prospective, multicenter, single-arm, open-label clinical study (ClinicalTrials.gov: NCT00915681). Randomization was not performed due to ethical and feasibility constraints related to disease rarity and established antidotal use of silibinin in Europe. The first ten cases received silibinin under Emergency IND authorizations (January 2007–March 2009). Open IND enrollment under the multicenter study protocol ran from July 2009 through October 2017. Sample size was determined by enrollment during the study period; no formal power calculation was performed.

### Enrollment and Eligibility

Patients were identified through a consultation hotline accessible via POISINDEX. Eligibility was confirmed by the Principal Investigator in consultation with referring physicians. Participating physicians signed commitment forms agreeing to protocol adherence and submission of complete inpatient medical records following hospitalization.

Patients ≥2 years of age were eligible with a prespecified diagnostic triad: (1) foraged mushroom ingestion; (2) onset of emesis or diarrhea 6–24 hours post-ingestion; and (3) rapidly rising transaminases. A protocol amendment excluded patients presenting with volume-unresponsive oliguric acute kidney injury (AKI). One patient under 24 months of age was enrolled with parental consent and FDA approval as a protocol exception. Laboratory confirmation of amatoxin exposure was not required; diagnosis relied on the characteristic clinical syndrome, as mycological and toxicological confirmation are unreliably available in practice.

### Treatment Protocol (Santa Cruz Protocol)

All protocol interventions were administered intravenously.

*Hydration:* Patients received an initial crystalloid bolus (4–7 liters of lactated Ringer’s or normal saline) to restore intravascular volume and re-establish urine output, followed by maintenance infusion of D5 0.45% normal saline at ≥200 mL/hr. Target urine output was ≥150 mL/hr in adults and 2–3 mL/kg/hr in children. Additional fluid boluses were administered for urine output <100 mL/hr or lactate elevation >3.0 mmol/L.

*Enterohepatic Circulation Suppression:* Patients were maintained nil per os (NPO) to prevent meal-triggered gallbladder contraction. Following a 2015 protocol amendment, octreotide was administered as a 200 µg bolus followed by continuous infusion at 50 µg/hr to pharmacologically inhibit gallbladder contraction and sphincter of Oddi relaxation. Both interventions continued until two consecutive INR measurements, obtained 6 hours apart, demonstrated improvement.

*Silibinin:* Intravenous silibinin dihemisuccinate (Legalon SIL, Madaus GmbH/Rottapharm|Mylan, now Viatris) was administered as a 5 mg/kg loading dose followed by continuous infusion of 20 mg/kg/day until INR normalized (≤1.2).

### Protocol Amendments

In 2010, physician commitment forms were introduced to ensure protocol adherence and complete medical record recovery. In 2015, serial lactate monitoring was added to provide real-time assessment of elimination capacity; octreotide was incorporated to pharmacologically suppress enterohepatic circulation during the interval before silibinin availability; and written treatment guidelines were distributed at enrollment.

### Pre-Enrollment Treatments

Prior to enrollment, some patients received N-acetylcysteine (76%), activated charcoal (54%), or penicillin G (23%) at the discretion of treating physicians. These interventions were not part of the study protocol.

### Data Collection

Clinical and laboratory data were abstracted from complete inpatient medical records by study personnel. Missing data were <5% for all measured variables.

### Outcomes

The primary outcome was transplant-free recovery (TFR), defined as survival to hospital discharge without liver transplantation. We use “recovery” rather than the conventional “survival” because patients achieving this endpoint demonstrated complete restoration of hepatic function and returned to baseline health without chronic sequelae—an outcome distinct from transplant-free survival in other acute liver failure etiologies where residual dysfunction may persist. Death and transplantation were objective events ascertained from medical records.

Secondary outcomes included: (1) death; (2) liver transplantation; (3) time from silibinin initiation to first INR reduction; (4) time to INR normalization; and (5) hospital length of stay.

### Analysis Populations

*Safety Population (N=99):* All patients receiving ≥1 dose of silibinin.

*Protocol-Adherent Population (n=86):* A protocol-adherent population was defined based on adherence to key protocol components required to achieve therapeutic elimination targets. Classification was determined from contemporaneously documented treatment orders and timing parameters, assessed independently of clinical outcomes. Patients were excluded from the protocol-adherent population for: 1) presentation with volume-unresponsive oliguric AKI; 2) delayed silibinin initiation (>120 hours post-ingestion) or incomplete silibinin delivery (<24 hours of protocol-dose infusion; failure to achieve ≥20 mg/kg/day for ≥24 hours); or 3) failure to achieve prespecified hydration targets due to inadequate presentation volume replacement (2 liters) or sustained withdrawal of maintenance intravenous hydration to <150 mL/hr. Some patients met more than one exclusion criterion.

*Protocol-Adherent Population (n=86):* A protocol-adherent population was defined based on adherence to key protocol components required to achieve therapeutic elimination targets. Classification was determined from documented treatment orders and timing parameters, recorded prospectively and assessed independently of clinical outcomes. Patients were excluded from the protocol-adherent population for: 1) presentation with volume-unresponsive oliguric AKI; 2) delayed silibinin initiation (>120 hours post-ingestion) or incomplete total dose (20 mg/kg); or 3) failure to achieve prespecified hydration targets due to inadequate presentation volume replacement (2 liters) or sustained withdrawal of maintenance intravenous hydration to <150 mL/hr. Some patients met more than one exclusion criterion.

### Statistical Analysis

*Primary Analysis:* TFR rates with exact binomial 95% confidence intervals were calculated for the Safety and Protocol-Adherent populations.

*Severity Assessment:* Baseline severity was characterized using the Model for End-Stage Liver Disease (MELD) score, calculated as: 3.78×ln(bilirubin mg/dL) + 11.2×ln(INR) + 9.57×ln(creatinine mg/dL) + 6.43. MELD scores were not capped at 40 to preserve the full severity distribution. The relationship between MELD score and outcome was assessed by logistic regression separately in each analysis population.

*Univariable Analysis:* Candidate predictors were evaluated individually for association with TFR using logistic regression (continuous variables) or Fisher’s exact test (categorical variables). Variables assessed included: age, sex, symptom latency (ingestion-to-symptom interval), presentation timing (ingestion-to-first-laboratory interval), peak laboratory values (ALT, AST, bilirubin, INR, creatinine, lactate, hemoglobin), treatment timing (ingestion-to-silibinin interval), total silibinin dose, hydration status, octreotide use, N-acetylcysteine use, and activated charcoal use.

*Multivariable Analysis:* Variables with P0.10 in univariable analysis were entered into multivariable logistic regression using stepwise selection (entry P0.10, retention P0.05). Model discrimination was assessed by the c-statistic (area under the receiver operating characteristic curve). Given the limited number of outcome events (n=12 poor outcomes), the final model was restricted to ≤2 predictors to avoid overfitting.

*Secondary Analyses:* Time from silibinin initiation to INR recovery was analyzed using linear regression with candidate predictors including symptom latency, octreotide use, and baseline INR. Kaplan-Meier methods were used to estimate survival distributions among patients with poor outcomes.

*Sensitivity Analyses:* To assess the robustness of findings, analyses were repeated: (1) excluding the 10 Emergency IND patients enrolled before the formal protocol; (2) stratified by MELD score (20 vs. ≥20); and (3) stratified by sex to address the observed imbalance in poor outcomes.

*Missing Data:* Complete case analysis was used. Missing values were not imputed given the low rate of missingness (5% for all variables).

All analyses were performed using SAS-JMP (SAS Institute, Cary, NC). Two-sided P<0.05 was considered statistically significant.

## RESULTS

### Participant Flow and Baseline Characteristics

Of 118 patients screened, 99 met eligibility criteria and received ≥1 dose of silibinin, constituting the Safety Population (Figure 1). Thirteen patients were excluded from the protocol-adherent population due to volume-unresponsive oliguric AKI at presentation (n=1), delayed silibinin initiation (>120 hours) or incomplete dosing (<20 mg/kg total; n=6), or failure to achieve hydration targets (n=8); some patients met multiple exclusion criteria. No patients were lost to follow-up. The protocol-adherent population was defined a priori to represent receipt of the elimination-preserving components necessary to test the mechanistic hypothesis that outcomes depend on maintained renal elimination capacity.

Mean age was 52.5 years (range 1.6–94); 42% were female. Median ingestion-to-symptom interval was 12 hours (IQR 8–24), consistent with amatoxin poisoning. Median time from ingestion to first laboratory evaluation (a surrogate for presentation timing) was 41.5 hours (IQR 28–61). Mycological species identification was available in 15 cases (15%), all confirming amatoxin-containing species.

### Baseline Severity

Patients developed severe hepatic dysfunction. In the Safety Population, ALT exceeded 2000 U/L in 78 patients (79%), and INR was ≥1.5 in 69 (70%), ≥2.0 in 47 (47%), and ≥4.0 in 21 (21%). Peak INR reached 14.9. Median peak MELD score was 22.3 (IQR 15–32); 17 patients (18%) had MELD ≥40.

The protocol-adherent population had similar baseline transaminase elevation (ALT >2000 U/L in 77%; INR ≥1.5 in 69%; peak INR 14.9) and modestly lower markers of hepatic dysfunction: median peak bilirubin 3.1 vs. 3.3 mg/dL (P=0.41), median peak INR 1.9 vs. 2.0 (P=0.38), and median MELD 20.2 vs. 22.3 (P=0.29). Median peak lactate was 3.0 mmol/L in the protocol-adherent population versus 3.5 mmol/L overall (n=48 with lactate measured; P=0.18). These differences were not statistically significant, indicating substantial severity overlap between populations.

### Primary Outcome: Transplant-Free Recovery

In the Safety Population (N=99), transplant-free recovery occurred in 87 patients (88.0%; 95% CI 79.8–93.6), liver transplantation in 6 (6.1%), and death in 6 (6.1%). Median hospital length of stay among survivors was 6 days (IQR 5–8).

In the protocol-adherent population (n=86), transplant-free recovery occurred in 85 patients (98.8%; 95% CI 93.7–100.0), with 1 transplantation and no deaths. The single transplant recipient presented with lactate 7.0 mmol/L that never corrected despite sustained hydration and azotemia resolution, suggesting hepatic injury too advanced for transplant-free recovery; sustained hydration nonetheless prevented oliguric AKI and hepatic encephalopathy, permitting stabilization until transplantation.

### MELD Score and Outcome

In the Safety Population, MELD score strongly predicted outcome: each 10-point MELD increase was associated with significantly lower transplant-free recovery probability (OR 0.35; 95% CI 0.19–0.62; P<0.001). The logistic model showed excellent discrimination (c-statistic 0.89). Among patients with MELD ≤20 (n=45), transplant-free recovery was 97.8% (44/45); among those with MELD 20–39 (n=34), 88.2% (30/34); among those with MELD ≥40 (n=17), 58.8% (10/17).

In striking contrast, the protocol-adherent population showed no significant relationship between MELD score and outcome (OR 0.72 per 10-point increase; 95% CI 0.31–1.67; P=0.44; c-statistic 0.62). Transplant-free recovery exceeded 95% across all MELD strata: 100% (39/39) for MELD ≤20, 97.8% (44/45) for MELD 20–39, and 100% (6/6) for MELD ≥40. This attenuation of the expected severity-outcome relationship suggests that protocol adherence—specifically, sustained hydration preserving elimination capacity—modified the natural history of severe amatoxin poisoning.

### Sex and Outcome

Among the 12 patients with poor outcomes (death or transplantation), 9 (75%) were female, despite females comprising 42% of the Safety Population (P=0.03 by Fisher’s exact test). In univariable analysis, female sex was associated with increased risk of poor outcome (OR 4.1; 95% CI 1.1–16.1; P=0.04). However, this association was attenuated and no longer significant after adjustment for MELD score (adjusted OR 2.8; 95% CI 0.6–12.4; P=0.18), suggesting that the sex imbalance was partly mediated by greater disease severity in women—potentially reflecting higher weight-adjusted toxin exposure.

### Predictors of Outcome: Univariable Analysis

Variables significantly associated with transplant-free recovery in univariable analysis included: sustained hydration (OR 28.4; 95% CI 6.8–119; P<0.001), ingestion-to-silibinin interval (OR 0.96 per hour; 95% CI 0.93–0.99; P=0.003), peak INR (OR 0.51 per unit; 95% CI 0.36–0.72; P<0.001), peak lactate (OR 0.42 per mmol/L; 95% CI 0.22–0.79; P=0.007), peak bilirubin (OR 0.83 per mg/dL; 95% CI 0.73–0.94; P=0.004), and peak creatinine (OR 0.63 per mg/dL; 95% CI 0.41–0.97; P=0.04).

Variables not significantly associated with transplant-free recovery included: age (P=0.31), symptom latency (P=0.52), peak ALT (P=0.14), N-acetylcysteine use (P=0.12), activated charcoal use (P=0.19), and total silibinin dose (P=0.28).

### Predictors of Outcome: Multivariable Analysis

Given 12 outcome events, the multivariable model was limited to 2 predictors to avoid overfitting. Stepwise selection identified sustained hydration and ingestion-to-silibinin interval as independent predictors of transplant-free recovery:

- Sustained hydration: OR 24.1 (95% CI 5.2–112); P<0.001
- Ingestion-to-silibinin interval: OR 0.95 per hour (95% CI 0.92–0.99); P=0.003

The model showed good discrimination (c-statistic 0.91) and fit (Hosmer-Lemeshow P=0.68). When MELD score was forced into the model, sustained hydration remained the dominant predictor (P<0.001), while MELD was no longer significant (P=0.14)—indicating that hydration status captured prognostic information beyond baseline severity alone.

### Octreotide and Time to INR Recovery

Among patients achieving transplant-free recovery with documented INR recovery timing (n=64), octreotide administration (n=35 in this subgroup; 38 total) was associated with shorter time from silibinin initiation to first INR improvement: mean 20.8 hours (SE 4.0) with octreotide versus 32.9 hours (SE 3.4) without (difference 12.1 hours; 95% CI 1.0–23.2; P=0.033). In multivariable analysis adjusting for symptom latency, octreotide remained independently associated with faster INR recovery (P=0.033), as did longer symptom latency (P=0.016)—the latter consistent with smaller ingestion and greater pre-presentation toxin elimination in patients with longer latency.

### Safety

Treatment-emergent adverse events occurred in 24 patients (24%), totaling 29 events. No cases of fluid overload, pulmonary edema, or heart failure occurred during protocol-specified hydration. Infusion-related reactions (warmth, flushing) occurred in 8 patients (8%) and resolved with rate adjustment; no serious adverse events were attributed to silibinin, or to octreotide (0/38 patients).

Two patients (2%) developed biopsy-confirmed acute tubular necrosis following hepatic recovery; one required temporary hemodialysis, and both subsequently recovered baseline renal function. These events occurred during the recovery phase, consistent with delayed manifestation of nephrotoxic injury sustained during the pre-presentation interval rather than treatment-related toxicity.

### Characteristics of Poor-Outcome Patients

All 12 patients with poor outcomes shared a common clinical trajectory: persistent lactate elevation (>3.0 mmol/L) that failed to correct with volume resuscitation, followed by oliguric AKI and subsequent multisystem failure. Among the 6 deaths, median time from ingestion to death was 179 hours (95% CI 115–281).

Eight of 12 poor-outcome patients had documented hydration restriction or inadequate volume resuscitation—either insufficient presentation bolus (<2 liters) or subsequent maintenance hydration reduction, typically following transfer to transplant centers where conservative fluid management is standard practice for other ALF etiologies. In several cases, renal function and/or lactate initially corrected with volume replacement at community hospitals, then rebounded after hydration withdrawal/reduction upon transplant center admission (Patients 1, 2, 3, 76, 188; Supplementary Appendix).

One patient presented with established oliguric AKI and made no urine despite fluid challenge, representing elimination failure at presentation. One patient presented 126 hours post-ingestion with INR 14 and uncorrectable lactate, reflecting injury too advanced for the elimination-based strategy. The remaining poor-outcome patients demonstrated the reproducible sequence: hydration reduction → lactate rebound → oliguric AKI → multisystem collapse.

### Biliary Drainage

Seven patients underwent biliary drainage procedures (5 ERCP with nasobiliary drain placement, 2 percutaneous cholecystostomy). One patient developed oliguric AKI following transplant-center hydration reduction but recovered completely after undergoing ERCP-guided nasobiliary drainage, with INR improvement within 24 hours—a recovery timeframe similar to that observed with silibinin, and consistent with the hypothesis that both interventions prevent hepatocellular re-exposure to sequestered amatoxin. The limited number of drainage procedures precludes formal statistical comparison, but these cases provide proof-of-concept for biliary drainage as an alternative when silibinin is unavailable or elimination capacity is compromised.

## DISCUSSION

In this prospective multicenter single-arm clinical trial, progression to multisystem failure following amatoxin mushroom ingestion was associated with loss of toxin elimination capacity rather than inexorable primary hepatic injury alone. Preservation of renal clearance, EHC suppression, and subsequent silibinin administration were associated with near-universal transplant-free recovery despite severe hepatic dysfunction, while largely eliminating the expected MELD severity–outcome relationship in the protocol-adherent population. These findings resolve the longstanding clinical paradox of deterioration during apparent recovery and establish a toxicokinetic framework for understanding both recovery and collapse—one in which fluid and dietary management decisions, rather than ingested dose or baseline severity alone, largely determine outcomes.

### The Kinetic Balance Between Clearance and Uptake

Under intact renal perfusion, >80% of the amatoxin absorbed escapes first-pass hepatic extraction,^11–15,21,22^ entering the systemic bloodstream for rapid glomerular filtration (>80% urinary recovery within 6 hours experimentally in dogs);^11–15^ sinusoidal OATP1B3 captures the remainder. This kinetic balance governs clinical trajectory. When renal clearance fails—most commonly from progression of subclinical tubular injury to established oliguric AKI—the fraction no longer cleared is distributed to the liver with each cardiac cycle,^30^ directly via the hepatic artery, and indirectly by the portal vein (draining the entire splanchnic circulation).^35^ We term this phenomenon *arteriohepatic uptake*—hepatic delivery from the systemic circulation—to distinguish it from first-pass extraction and EHC: it represents systemic distribution to the liver that occurs when the renal clearance pathway is compromised.^30^ The arteriohepatic surge in hepatocellular exposure precipitates the abrupt transition to multisystem failure characteristic of fatal amatoxin poisoning.

This kinetic mismatch explains how loss of clearance secondary to oliguric AKI can trigger the hepatic uptake surge that precipitates multisystem failure, why even small ingestions may become lethal when elimination fails,^5,6^ and why risk rises the longer presentation is delayed as nephrotoxic exposure accumulates—underscoring the singular exigency for rapid, generous volume replacement at presentation. This framework inverts the conventional understanding: kidney failure does not result from hepatic collapse but rather precipitates it.

### The Toxicokinetic Cascade

The clinical course reflects sequential phases of toxin distribution shaped by physiologic events and clinical decisions. Following ingestion, rapid absorption from stomach and intestine delivers toxin to the portal circulation; first-pass hepatic extraction is limited (<20%), with the majority entering the systemic circulation for renal clearance during presymptomatic latency.^11–15^ Hepatocytes excrete unbound toxin into bile, which accumulates in the gallbladder.

During presymptomatic latency, meal-triggered EHC serves a beneficial function: reintroduced toxin is predominantly cleared by functioning kidneys (>80%), a minority returning to exacerbate hepatotoxicity—a favorable trade-off under intact perfusion. The frequently cited figure of 60% enterohepatic recycling has no identifiable primary source and likely overstates the contribution when renal clearance is preserved. Smaller ingestions produce longer latency, allowing more elimination time but also more meal opportunity; larger ingestions have shorter latency, leaving less time for both.

This equilibrium deteriorates as the gastrointestinal phase unfolds. Enormous, easily underestimated fluid losses produce hypovolemia and prerenal AKI—marked hemoglobin elevation at presentation is typical. However, the hepatotoxic consequences are limited: most absorbed amatoxin has already undergone renal clearance. In the absence of oral intake, EHC remains quiescent;^19^ gallbladder accumulation isolates hepatocyte-excreted toxin from both systemic circulation and gut. Meanwhile, the prolonged interval from symptom onset to volume resuscitation (median >41 hours in this series) promotes concentration– and time-dependent nephrotoxicity during tubular stasis—injury that remains subclinical when perfusion is promptly restored and tubular washout maintained, but can progress to established oliguric AKI if hydration support is delayed, inadequate, reduced, or withdrawn.^20–22,36^

The critical vulnerability emerges during the pseudo-recovery phase, when two clinical decisions converge with devastating consequences. Meal resumption—ordered as hemodynamic and gastrointestinal symptoms resolve and appetite returns—triggers cholecystokinin-mediated gallbladder contraction and sphincter of Oddi relaxation, releasing sequestered amatoxin-laden bile back into the duodenum for renewed absorption and systemic reintroduction. Simultaneously, reduction or withdrawal of intravenous hydration—ordered as patients appear euvolemic—unmask subclinical tubular injury, compromising clearance precisely when new toxin burden appears.^37^ The favorable EHC trade-off that existed during latency now inverts catastrophically: with clearance compromised, the >80% that would have been eliminated is instead distributed to the liver.^30^ This convergence precipitates the arteriohepatic uptake surge that triggers multisystem collapse. The reproducible sequence observed in poor-outcome patients (hydration reduction → lactate rebound → oliguric AKI → multisystem failure) confirms this mechanism. Pseudo-recovery thus represents a vulnerable equilibrium dependent on continued toxin elimination rather than an unpredictable interval of occult decline. Fluid and dietary management decisions determine whether that equilibrium holds or collapses.

### Hydration as the Critical Modifiable Factor

Multivariable analysis identified sustained hydration as the strongest predictor of transplant-free survival. Ample presentation volume replacement reverses prerenal AKI, restores elimination, and—critically—prevents occult tubular injury from progressing to established oliguric AKI.^38^ This explains why the window for intervention is presentation: prompt volume restoration and sustained hydration suppress functional manifestation of subclinical injury and initiate healing;^12,20–22,38^ delay, under-resuscitation, or subsequent fluid withdrawal allows ATN to become established, setting the stage for the calamitous pseudo-recovery convergence. Following initial resuscitation with isotonic crystalloid, maintenance with hypotonic fluid (D5 0.45NS at ≥200 mL/hour) provided volume and free water support while reducing overload risk,^39^ sustaining urine output above 150 mL/hour to optimize ongoing elimination.^12,21,22^ Experimental and clinical data support this framework. Autopsy series consistently demonstrate ATN—direct tubular injury—rather than the functional renal failure of hepatorenal syndrome.^40,41^ In a porcine model, Thiel et al. demonstrated that sustained fluid support alone—without antidotal therapy—enabled spontaneous recovery in 4 of 5 landrace pigs receiving LD50 α-amanitin, while extending survival and delaying multisystem failure until 96 hours even in lethally intoxicated LD100-dosed animals.^42^ Enhanced hydration prevented oliguric AKI and improved outcomes in multiple Italian cohorts;^22,43,44^ implementation of mandatory multi-liter volume replacement at an Assamese university hospital reduced mortality from 44% to 14%.^5^ Published series consistently document oliguric AKI preceding severe hepatic deterioration.^7–10,22,40^ In this study, patients who did not receive sustained hydration experienced substantially worse outcomes despite similar initial severity.

This observation has important implications for tertiary transfer. Conservative fluid management—often used in other acute liver failure etiologies to mitigate cerebral edema^45,46^—may be counterproductive in amatoxin poisoning, where renal perfusion is essential to maintaining the dominant elimination pathway. In several poor-outcome cases, lactate rebound soon followed by oliguria developed within hours of reduced maintenance hydration after transfer, consistent with elimination failure rather than institutional capability (Supplementary Appendix). Native kidneys provide rapid clearance of circulating amatoxin that is not meaningfully replaced by extracorporeal support.^12,21,22,45^ Renal replacement therapy, frequently initiated for hyperammonemia management, does not achieve clinically meaningful amatoxin removal and—when coupled with reduced intravenous hydration—may further compromise renal perfusion.^45^ Accordingly, therapies or management decisions that predictably reduce effective renal perfusion—including hydration restriction or medications with hemodynamically consequential renal effects—may precipitate oliguric AKI and accelerate clinical deterioration.

### Enterohepatic Circulation and Gallbladder Sequestration

This study supports a crucial role for the gallbladder as the principal reservoir of uneliminated toxin. Hepatocytes rapidly excrete unbound amatoxin into bile,^11,13,15,17^ which accumulates in the gallbladder between meals. Meal-induced contraction releases amatoxin-laden bile into the intestine, enabling renewed absorption and hepatic uptake in a cycle that exacerbates hepatotoxicity until bile and toxin reaccumulate. Biochemical analysis by Leite et al. confirmed substantial amatoxin content in gallbladder aspirates obtained from patients several days post-ingestion,^18^ consistent with prolonged sequestration.

The clinical significance of EHC has been questioned based on a 2011 porcine study reporting declining biliary amanitin concentrations.^48^ However, the anesthesized animals received high-dose fentanyl (600 μg/hr), which potently inhibits cholecystokinin release and gallbladder contraction—pharmacologically suppressing the mechanism under investigation.^49^ Furthermore, common bile duct effluent rather than gallbladder contents was sampled, missing the primary sequestration compartment.^49,50^ Leite’s biochemical analysis of actual gallbladder aspirates resolved this apparent discrepancy.^18^

Augmenting fasting with octreotide (n=38, enrolled 2015–2017), which inhibits cholecystokinin-mediated gallbladder contraction and sphincter of Oddi relaxation,^51–53^ was associated with a 12-hour reduction in time to INR recovery, consistent with reduced hepatocellular re-exposure during the pre-silibinin interval. Octreotide thus serves as a pharmacological bridge, maintaining gallbladder sequestration until silibinin establishes hepatoprotection.

### Silibinin and the Therapeutic Window

Strong association between silibinin administration and transplant-free survival supports antidotal benefit beyond hydration alone and demonstrates a therapeutic window extending to 120 hours post-ingestion—more than doubling previous estimates.^26,28^ By blocking OATP1B3-mediated hepatic uptake, silibinin diverts sinusoidal amatoxin into the systemic circulation for renal clearance—a mechanism requiring functioning kidneys.

Serial lactates (available in 48 patients, predominantly after protocol amendment) provided early insight into elimination capacity: sustained correction preceded recovery, whereas persistent elevation predicted failure. Presentation lactate (2.0–8.0 mmol/L) corrected with volume replacement in 60% of patients, including eight with levels exceeding 4.0 mmol/L. Treatment failures were consistently preceded by oliguric AKI, suggesting silibinin efficacy depends on intact renal elimination. Persistent lactate elevation (>3.5 mmol/L beyond 12–24 hours of volume optimization) despite adequate urine output may identify patients in whom silibinin’s renal-dependent mechanism is unlikely to succeed. Serial lactate thus serves not only as a prognostic marker but as a potential surrogate for elimination capacity itself—correction reflecting restored clearance, persistent elevation signaling ongoing toxin redistribution or hepatic injury too advanced for silibinin to reverse.

### Gallbladder Drainage as a Resource-Independent Alternative

When silibinin is unavailable or likely to fail (persistent lactate elevation, oliguric AKI), gallbladder drainage provides a toxicokinetically sound therapeutic alternative,^15,17,50^ directly evacuating amatoxin-laden bile from its principal accumulation reservoir. Experimentally, beagles with surgical biliary diversion survived otherwise lethal amanitin exposure.^15,17^ Case reports,^4,54–65^ human and veterinary,^66,67^ consistently describe durable INR improvement within 24–36 hours following drainage—a timeframe similar to silibinin, consistent with ALF reversal when hepatocellular re-exposure is curtailed. One patient developed oliguric AKI after transplant-unit fluid restriction yet achieved full recovery following ERCP-guided nasobiliary drainage—the only such patient to escape death or transplantation.

Multiple approaches have been described, including needle aspiration, percutaneous or surgical cholecystostomy, and ERCP-guided nasobiliary drainage. Procedural risks (bile leak peritonitis, bleeding, and infection)^68,69^ must be weighed against outcome risks (liver transplantation, death) associated with limited elimination capacity. Importantly, drainage requires no specialized pharmaceuticals and can be performed in any facility with basic interventional or surgical capability. This mechanistic equivalence—both silibinin and drainage prevent hepatocellular re-exposure to sequestered amatoxin—explains the similar recovery timeframes and supports a unified toxicokinetic model.

### Conventional Interventions

Neither N-acetylcysteine nor activated charcoal was associated with transplant-free survival or accelerated INR recovery. N-acetylcysteine lacks toxicokinetic rationale^70^—amatoxin is not metabolized—demonstrated no benefit in animal models,^71^ and marginally improved survival in one adult non-acetaminophen ALF trial (P=.043),^72^ but not a pediatric;^73^ and neither included amatoxin patients).^69,70^ A transient INR-raising effect may confound prognostic assessment.^74,75^ Penicillin G, despite widespread use, showed no outcome association in two large multivariable analyses.^76,77^ Activated charcoal is often poorly tolerated, with recognized aspiration and ileus risk.^78^ Amatoxin’s polarity, high water-solubility, and rapid absorption make it a poor charcoal substrate; once absorption is complete, intestinal toxin burden remains low until meal-triggered EHC during pseudo-recovery, obviating the rationale for delayed or multi-dosing. In several cases, extensive adjunctive therapy coincided with inadequate volume replacement, illustrating how elimination-critical priorities can be displaced by interventions of unproven benefit.^79,80^

### Implications for Global Practice

Recent reports from Asia (including China,^81^ Thailand,^82^ Iran,^83^ and Turkey^84^), the Indian subcontinent,^41,85^ southern Africa,^86^ and the Americas suggest an underrecognized global burden,^87–89^ compounded by overseas expansion of *Amanita phalloides*—its unseen mycelia permeating the soil of potted tree exports.^90^ Fatal outcomes are not WHO-reportable; the true burden remains uncounted, particularly in subtropical regions where fatalities follow misidentification of look-alike edibles,^91^ and may be misclassified as endemic diarrheal illness.^92^

These considerations underscore the need for a resource-neutral management paradigm. The core protocol components—crystalloid hydration, fasting, and octreotide (a WHO essential medicine)—require no specialized infrastructure. Gallbladder drainage provides definitive intervention when silibinin is unavailable. This framework offers a physiologically grounded path to improving outcomes from tertiary transplant centers to the resource-limited facilities where most amatoxin poisoning fatalities occur.^92^

### Limitations

This study has limitations inherent to researching treatment for unpredictably occurring, rapidly fatal poisonings.^93,94^ Single-arm design precludes randomized comparison, and the protocol-adherent population analysis may be subject to selection bias. However, MELD-stratified analysis demonstrated substantial severity overlap between populations, and outcomes diverged sharply with hydration adherence even among high-MELD patients—supporting management-dependent rather than severity-dependent outcomes.

One protocol-adherent patient with advanced injury at presentation (INR 7.8, lactate persistently elevated despite enhanced hydration and creatinine correction) required transplantation; notably, neither hepatic encephalopathy nor oliguric AKI developed preoperatively, suggesting the protocol preserved physiologic stability even when hepatic reserve was insufficient for TFR. This case illustrates the limits of elimination-based therapy: it cannot regenerate a liver already destroyed but may prevent complications that would otherwise preclude successful transplantation.

Investigator involvement may have influenced adherence, although outcomes were objective and ascertainment relied on primary medical records. Laboratory confirmation was unavailable; diagnosis relied on a characteristic clinical syndrome. Protocol refinements during the study reflected evolving mechanistic understanding. The 3.5 mmol/L lactate threshold emerged from clinical observation rather than prospective validation.

### Conclusions

This prospective trial demonstrates that progression to multisystem failure in amatoxin mushroom poisoning is associated with loss of toxin elimination capacity rather than inexorable primary hepatic injury. The conventional assumption—that liver failure drives kidney failure—has the causal relationship inverted: it is oliguric kidney failure that precipitates hepatic collapse.

The findings establish a kinetic model in which preserved renal clearance and gallbladder sequestration prevent the surge of arteriohepatic uptake that precipitates collapse; pseudo-recovery represents a vulnerable equilibrium, and the convergence of meal resumption and hydration withdrawal triggers the fatal cascade. Clinical decisions—not disease biology alone—determine whether that equilibrium holds. The results support a toxicokinetic-based management strategy prioritizing preservation of elimination pathways: generous presentation volume replacement to restore renal perfusion and toxin clearance; sustained enhanced hydration to prevent progression from subclinical tubular injury to oliguric AKI and maintain elimination; fasting and octreotide to suppress enterohepatic circulation; and silibinin or gallbladder drainage to prevent hepatocellular re-exposure to sequestered toxin. This framework provides the first prospective evidence defining recovery determinants in amatoxin poisoning and offers a globally applicable approach to improving outcomes.

## DECLARATIONS

### Author contributions

Concept and design: Mitchell, Spyker, Robbins.

Acquisition, analysis, or interpretation of data: Spyker, Mitchell.

Drafting of the manuscript: Mitchell, Spyker, Robbins, Rumack.

Critical revision of the manuscript for important intellectual content: Mitchell, Spyker, Rumack. Statistical analysis: Spyker.

Mitchell and Spyker had full access to all data and take responsibility for the integrity of the data and the accuracy of the analyses. All authors reviewed and approved the final manuscript.

### Data availability

Data supporting the findings of this study are available in the Supplementary Materials. Additional de-identified data may be made available from the corresponding author upon reasonable request and subject to applicable institutional approvals and data-use requirements.

## Acknowledgments

We thank the Dominican Hospital Pharmacy staff for round-the-clock storage, packing, and shipment of study drug throughout the study period. We are grateful for guidance and support from colleagues at the U.S. Food and Drug Administration, including Sandra Saniga. We thank Manuel Ibarra and Paul Erhardt for their assistance in terming “arteriohepatic uptake.” We also acknowledge Heinz Faulstich, Thomas Zilker, Martin Langer, Larrimore Cummins, Massimo D’Amato, Joachim Maus, Michael Purzycki, Joe Veilleux, Donna Griebel Alan Buchwald, Kent Olson, Phool Konwar Deori, Jou-Fang Deng, Yuming Wang, Geng Jiawei, and Ma Xiuying.

We sincerely thank the participating clinicians and hospitals where patients were managed:

Aria Health Frankford Campus. Philadelphia, PA

Arroyo Grande Community Hospital, Arroyo Grande, CA

Beth Israel Deaconess Medical Center, Boston, MA

California Pacific Medical Center, San Francisco, CA

Case Western Reserve University Hospital, Cleveland, OH

Cleveland Clinic, Cleveland, OH

Dominican Hospital, Santa Cruz, CA

Duke University Medical Center, Durham, NC

Georgetown University Hospital, Washington, DC

Georgetown University Medical Center, Washington, DC

Highland Hospital, Oakland, CA

Holy Cross Hospital, Washington, DC

Hotel-Dieu Grace Hospital, Windsor, Ontario, CANADA

Indiana University Health Medical Center, Indianapolis, IN

John Muir Health-Walnut Creek Medical Center, Walnut Creek, CA

Johns Hopkins Hospital, Baltimore, MD

Kaiser Permanente Medical Center, Roseville, CA

Kaiser Permanente Oakland, Oakland, CA

Kaiser Permanente Sacramento, Sacramento, CA

Kaiser Permanente San Jose, San Jose, CA

Kaiser Permanente Santa Rosa Medical Center, Santa Rosa, CA

Marin General Hospital. Greenbrae, CA

Morristown Medical Center, Morristown, NJ

Natividad Medical Center, Salinas, CA

Oregon Health & Science University, Portland, OR

Reston Hospital Center, Reston, VA

Royal Jubilee Hospital, Victoria, Vancouver, CANADA

Rush University Medical Center, Chicago, IL

St. Francis Hospital and Medical Center, Hartford, CT

St. Joseph Medical Center, Stockton, CA

St. Peter Hospital-Albany, Albany, NY

St. Elizabeth Youngstown Hospital, Youngstown, OH

Strong Memorial Hospital of the University of Rochester, Rochester, NY

Swedish Medical Center, Seattle, WA

Thomas Jefferson University Hospital, Philadelphia, PA

University of California, San Francisco, San Francisco, CA

University of Connecticut Health Center, Farmington, CT

University of Iowa Hospital, Iowa City, IA

University of Maryland Medical Center, Baltimore, MD

University of Medicine and Dentistry of New Jersey, Newark, NJ

University of Washington Medical Center, Seattle, WA

University Hospital, Newark, NJ

Virtua Our Lady of Lourdes Hospital, Camden, NJ

Yale New Haven Hospital, New Haven, CT.

Correspondence to: S. Todd Mitchell, MD, MPH.

## Tables and Figures

**Table 1.**
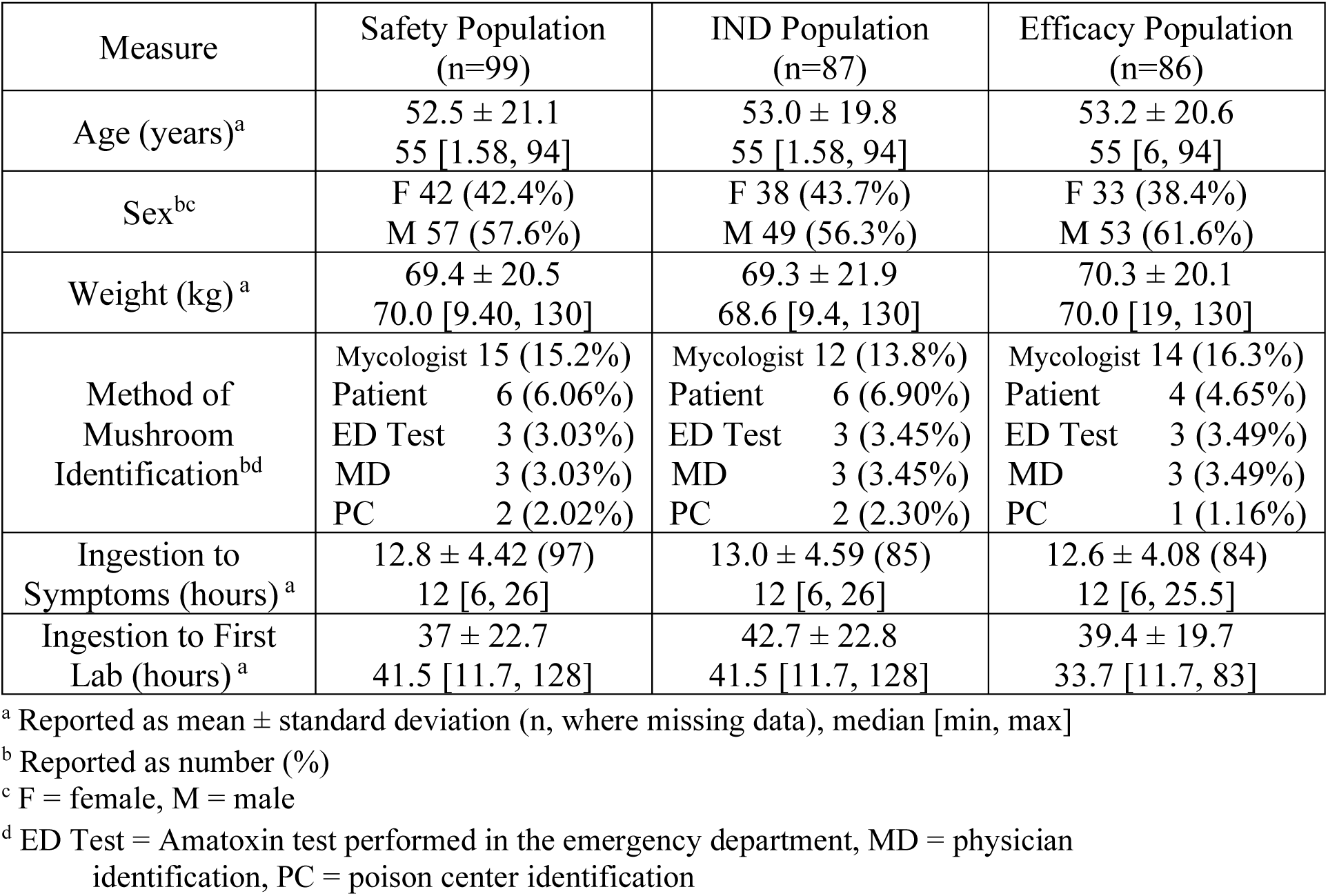
Description of Patients by Population.

| Measure | Safety Population<br>(n=99) | IND Population<br>(n=87) | Efficacy Population<br>(n=86) |
| --- | --- | --- | --- |
| Age (years)<sup>a</sup> | 52.5 ± 21.1<br>55 [1.58, 94] | 53.0 ± 19.8<br>55 [1.58, 94] | 53.2 ± 20.6<br>55 [6, 94] |
| Sex<sup>bc</sup> | F 42 (42.4%)<br>M 57 (57.6%) | F 38 (43.7%)<br>M 49 (56.3%) | F 33 (38.4%)<br>M 53 (61.6%) |
| Weight (kg)<sup>a</sup> | 69.4 ± 20.5<br>70.0 [9.40, 130] | 69.3 ± 21.9<br>68.6 [9.4, 130] | 70.3 ± 20.1<br>70.0 [19, 130] |
| Method of<br>Mushroom<br>Identification<sup>bd</sup> | Mycologist 15 (15.2%)<br>Patient 6 (6.06%)<br>ED Test 3 (3.03%)<br>MD 3 (3.03%)<br>PC 2 (2.02%) | Mycologist 12 (13.8%)<br>Patient 6 (6.90%)<br>ED Test 3 (3.45%)<br>MD 3 (3.45%)<br>PC 2 (2.30%) | Mycologist 14 (16.3%)<br>Patient 4 (4.65%)<br>ED Test 3 (3.49%)<br>MD 3 (3.49%)<br>PC 1 (1.16%) |
| Ingestion to<br>Symptoms (hours)<sup>a</sup> | 12.8 ± 4.42 (97)<br>12 [6, 26] | 13.0 ± 4.59 (85)<br>12 [6, 26] | 12.6 ± 4.08 (84)<br>12 [6, 25.5] |
| Ingestion to First<br>Lab (hours)<sup>a</sup> | 37 ± 22.7<br>41.5 [11.7, 128] | 42.7 ± 22.8<br>41.5 [11.7, 128] | 39.4 ± 19.7<br>33.7 [11.7, 83] |

<sup>a</sup> Reported as mean ± standard deviation (n, where missing data), median [min, max]
<sup>b</sup> Reported as number (%)
<sup>c</sup> F = female, M = male
<sup>d</sup> ED Test = Amatoxin test performed in the emergency department, MD = physician identification, PC = poison center identification

**Table 2.** Severity and Treatment by Population.

| Measure | Safety Population<br>(n=99) | IND Population<br>(n=87) | Efficacy Population<br>(n=86) |
| --- | --- | --- | --- |
| Max hemoglobin<br>(g/dL)<sup>a</sup> | 15.3 ± 2.15 (95)<br>15.1 [10.9, 21.4] | 15.3 ± 2.21 (87)<br>15.1 [10.9, 21.4] | 15.2 ± 2.00 (82)<br>15.1 [11.8, 21.4] |
| Max BUN<br>(mg/dL)<sup>a</sup> | 25.3 ± 14.4 (96)<br>21 [4, 64] | 25.2 ± 14.8 (87)<br>21 [4, 64] | 24.4 ± 14.0 (83)<br>21 [4, 64] |
| Max creatinine<br>(mg/dL)<sup>a</sup> | 1.57 ± 1.38 (96)<br>1.01 [0.31, 8.83] | 1.55 ± 1.37 (87)<br>1.0 [0.31, 8.83] | 1.43 ± 1.33 (83)<br>0.98 [0.31, 8.83] |
| Max AST<br>(U/L)<br>Num (%) >2000<sup>ab</sup> | 4467 ± 4255 (99)<br>3274 [15, 19972]<br>66 (66.7%) | 4361 ± 3941 (87)<br>3291 [15, 18117]<br>58 (66.7%) | 4065 ± 3911 (86)<br>3177 [15, 18117]<br>55 (64%) |
| Max ALT<br>(U/L)<br>Number (%) >2000<sup>a</sup> | 4872 ± 4011 (99)<br>4405 [31, 27117]<br>78 (78.8%) | 4729 ± 3802 (87)<br>4405 [31, 27117]<br>69 (79.3%) | 4621 ± 4111 (86)<br>3939 [31, 27117]<br>66 (76.7%) |
| Max total<br>bilirubin (mg/dL)<sup>a</sup> | 5.67 ± 5.28 (97)<br>3.32 [0.6, 25.8] | 5.50 ± 5.22 (87)<br>3.2 [0.6, 25.8] | 4.75 ± 4.31 (84)<br>3.1 [0.6, 19.6] |
| Max international<br>normalized ratio<sup>a</sup><br>Number (%) >1.5<sup>c</sup><br>Number (%) >2 | 3.26 ± 3.15 (99)<br>2.0 [1.0, 14.9]<br>69 (69.7%)<br>47 (47.5%) | 3.24 ± 3.06 (87)<br>2.0 [1.0, 14.1]<br>60 (69.0%)<br>42 (48.3%) | 2.45 ± 1.92 (86)<br>1.9 [1.0, 14.9]<br>56 (65.1%)<br>35 (40.7%) |
| Max Lactate<br>(mmol/L)<sup>ad</sup> | 5.00 ± 4.39 (48)<br>3.5 [1, 26] | 5.00 ± 4.39 (48)<br>3.5 [1, 26] | 3.41 ± 1.72 (37)<br>3.0 [1, 8] |
| MELD score<sup>ae</sup><br>Number (%) ≥40 | 24.8 ± 12.6 (96)<br>22.3 [6.71, 58.8]<br>17 (17.7%) | 24.6 ± 12.5 (87)<br>21.8 [6.71, 58.8]<br>15 (17.2%) | 21.7 ± 9.72 (83)<br>20.2 [6.71, 50.6]<br>6 (7.23%) |
| Ingestion to SIL<br>Start (hours)<sup>a</sup> | 65.5 ± 23.8 (79)<br>63.8 [15, 150] | 64.5 ± 25.1 (87)<br>61.7 [15, 150] | 63.8 ± 20.3 (86)<br>64.4 [19, 118] |
| SIL Dose<br>(mg/kg)<sup>a</sup> | 65.3 ± 36.1 (99)<br>61.1 [5.0, 221] | 67.0 ± 36.4 (87)<br>61.8 [15.0, 221] | 63.7 ± 31.3 (86)<br>60.7 [24.6, 182] |
| Duration of SIL<br>Treatment (days)<sup>af</sup> | 2.77 ± 1.64 (99)<br>2.47 [0.0271, 10.5] | 2.83 ± 1.66 (87)<br>2.51 [0.027, 10.5] | 2.67 ± 1.39 (86)<br>2.46 [0.027, 8.46] |

<sup>a</sup> Reported as mean ± standard deviation (number), median [min, max]
<sup>b</sup> ALT >2000 have defined “severe” amatoxin poisoning in several retrospective series
<sup>c</sup> INR >1.5 is a commonly accepted threshold for defining FHF
<sup>d</sup> Serum lactate data were not measured on patients enrolled early in the study
<sup>e</sup> MELD (Model for End-Stage Liver Disease) score = $3.78 \cdot \ln(\text{Bili}) + 11.2 \cdot \ln(\text{INR}) + 9.57 \cdot \ln(\text{Cr}) + 6.43$
<sup>f</sup> Includes the 18-30 hours typically required to deliver SIL from shipping depots

**Table 3.** Outcomes by Study Population.

| Measure | Safety Population<br>(n=99) | IND Population<br>(n=87) | Efficacy Population<br>(n=86) |
| --- | --- | --- | --- |
| Outcome<sup>ab</sup> | Death 6 (6.06%)<br>LT 6 (6.06%)<br>TFR 87 (87.9%) | Death 5 (5.7%)<br>LT 6 (6.9%)<br>TFR 76 (87.4%) | Death 0 (0%)<br>LT 1 (1.2%)<br>TFR 85 (98.8%) |
| Outcome<br>TFR mean<br>[95% CI]<sup>ac</sup> | Death/LT 12 (12.1%)<br>TFR 87 (87.9%)<br>[79.8%, 93.6%] | Death/LT 11 (12.6%)<br>TFR 76 (87.4%)<br>[78.5%, 93.5%] | Death/LT 1 (1.2%)<br>TFR 85 (98.8%)<br>[93.7%, 100%] |
| TFR Hospital<br>LOS Days<sup>de</sup> | 6.55 ± 2.57 (87)<br>6 [3, 16] | 6.50 ± 2.63 (76)<br>6 [3, 16] | 6.48 ± 2.51 (85)<br>6 [3, 16] |

<sup>a</sup> reported as number (%)
<sup>b</sup> LT = liver transplant, TFR = Transplant-free recovery
<sup>c</sup> Death/LT = death or liver transplant
<sup>d</sup> reported as mean ± standard deviation (n), median [min, max]
<sup>e</sup> Hospital days = Discharge Date – Admission date + 1

**Figure 1.**
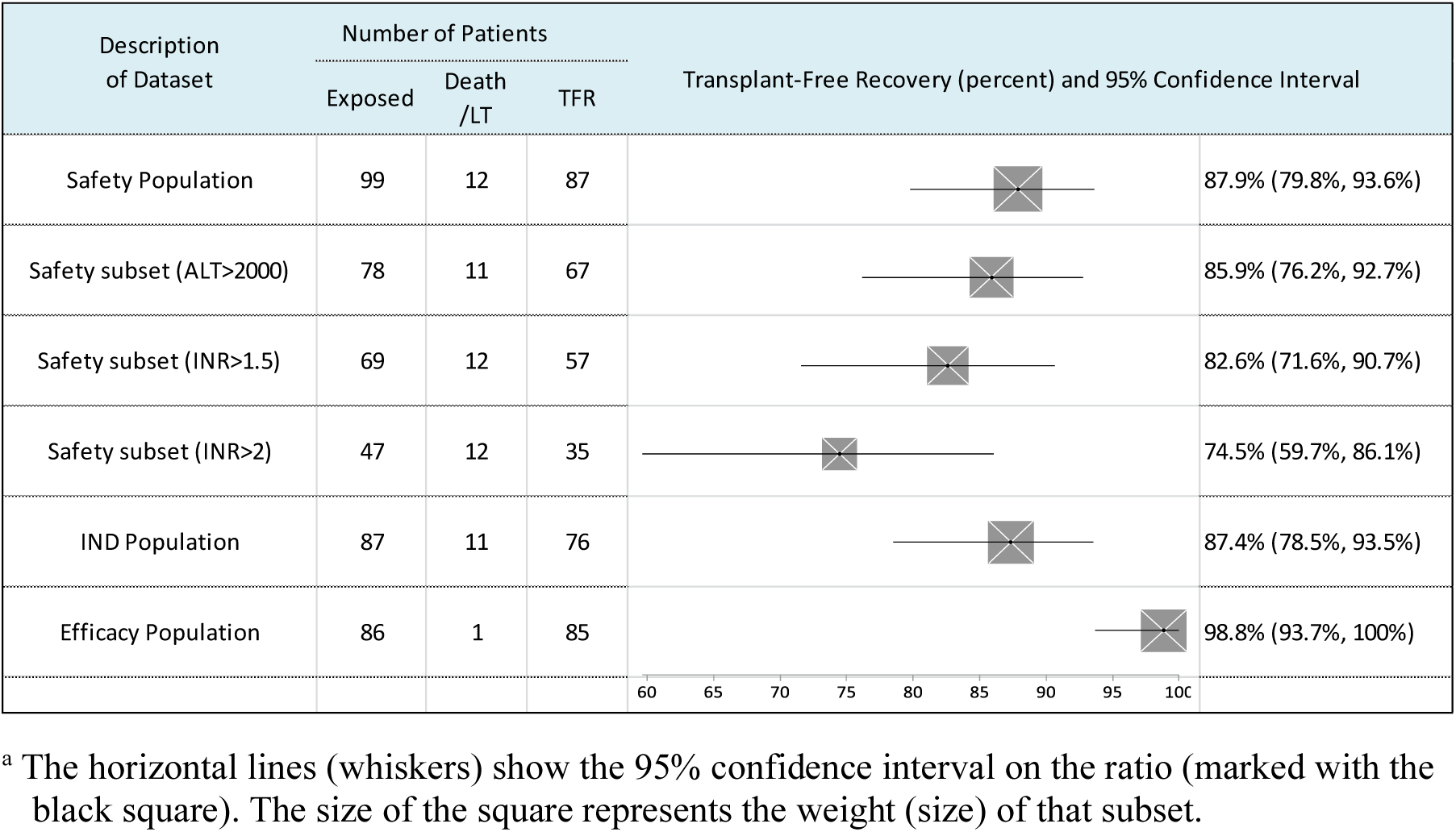
Subset Comparisons for Transplant-Free Recovery. Forest Plot^a^ comparing the 3 study populations and 3 severity subsets of the Safety Population

**Figure 2.**
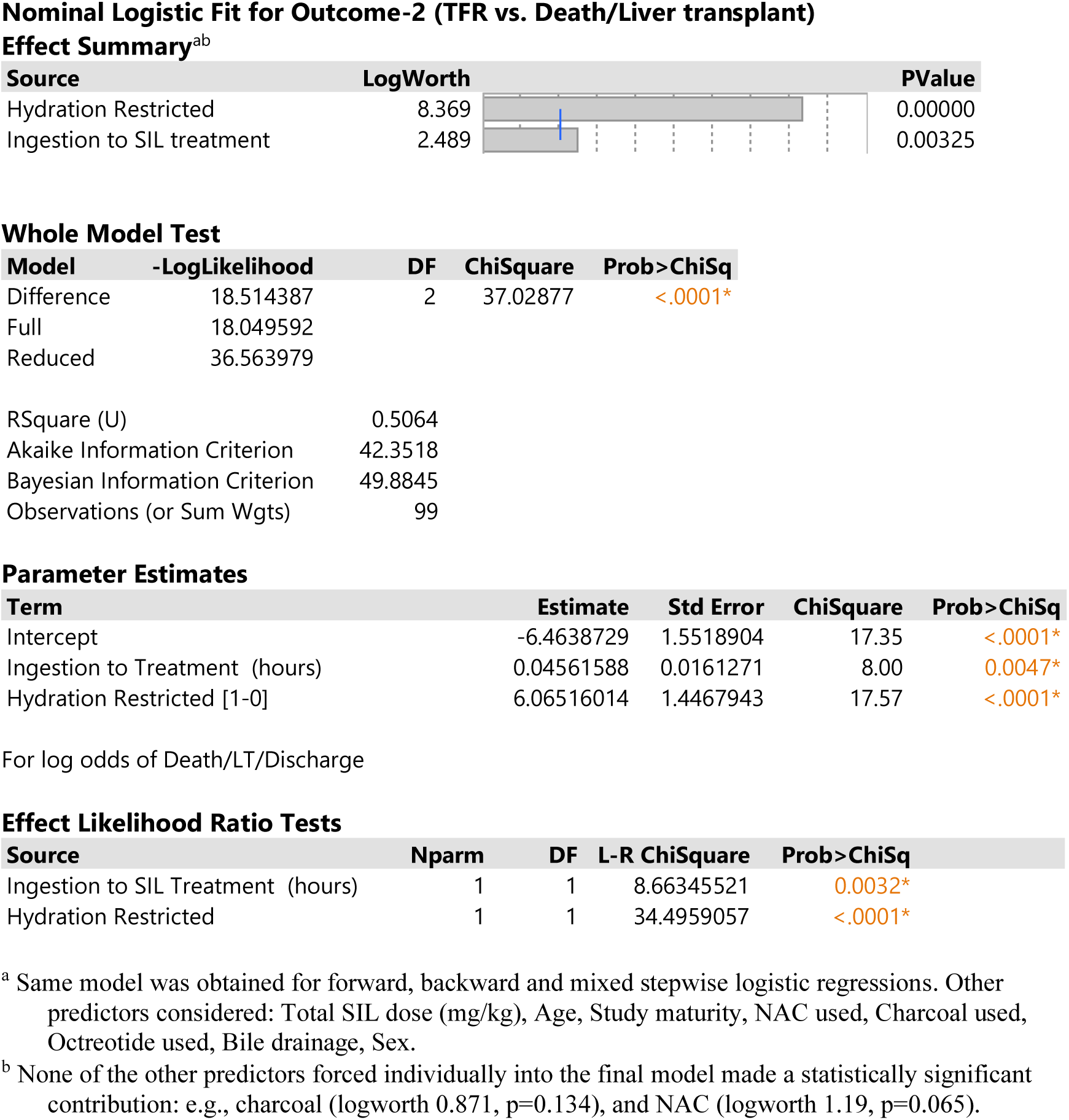
Multivariate Predictor of Outcome (TFR vs. Death/LT) Safety Population (n=99)

**Figure 3.**
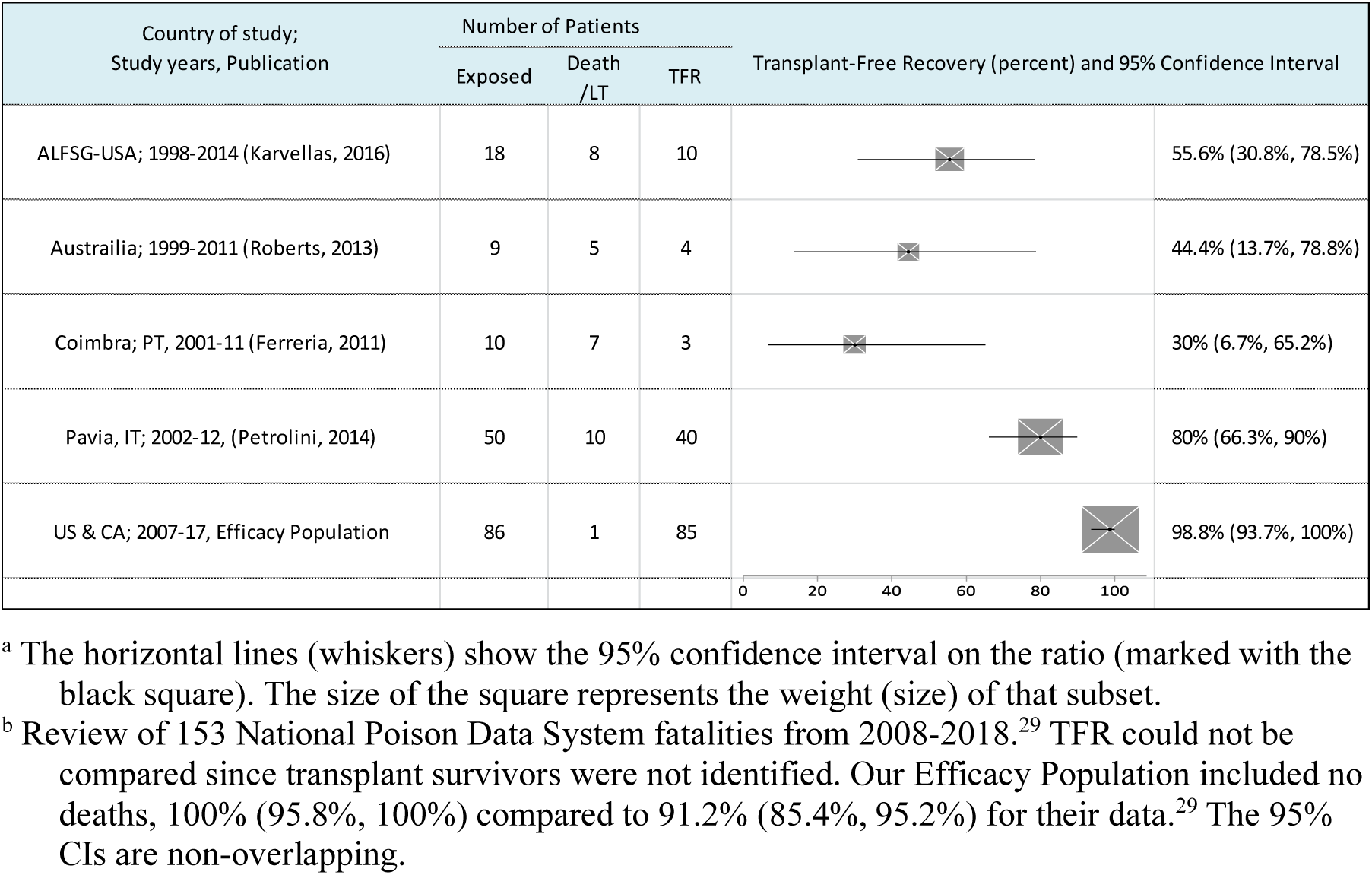
Transplant-Free Recovery Comparisons by Study. Forest Plot^a^ comparing the 4 published study populations^b^ to Efficacy Population

**Table 4.** Resolution of Clinical Enigmas in Amatoxin Poisoning Through Toxicokinetic Mechanisms.

| Clinical Enigma | Traditional Understanding | Toxicokinetic Resolution | Supporting Evidence |
| --- | --- | --- | --- |
| Unpredictable deterioration during apparent recovery | Occult hepatic decline during "pseudo-recovery" | Pseudo-recovery represents conditional equilibrium dependent on continued elimination; convergence of meal resumption and hydration withdrawal triggers arteriohepatic uptake surge | Reproducible sequence in poor-outcome patients (this trial) |
| Small ingestions sometimes fatal; large ingestions sometimes survived | Toxicity assumed to be strictly dose-dependent | Elimination capacity, not ingested dose, determines outcome; oliguric AKI precipitates redistribution regardless of initial dose | MELD-outcome relationship eliminated in protocol-adherent population (this trial) |
| Oliguric AKI consistently precedes hepatic collapse | Assumed to reflect hepatorenal syndrome (liver failure → kidney failure) | Causal relationship inverted: kidney failure precipitates hepatic collapse by terminating clearance and triggering arteriohepatic redistribution | ATN (not hepatorenal) on autopsy histopathology; temporal sequence documented (this trial) |
| Silibinin failures despite early administration | Unexplained treatment resistance | Silibinin efficacy requires intact renal elimination to clear diverted toxin; oliguric AKI renders hepatic protection insufficient | All treatment failures preceded by oliguric AKI (this trial) |
| Similar recovery timeframes with silibinin or biliary drainage | Coincidental or unexplained | Mechanistic equivalence: both interventions prevent hepatocellular re-exposure to gallbladder-sequestered amatoxin | Case reports; biochemical confirmation of gallbladder sequestration (Leite et al.) |
| Transplant-center outcomes no better than resource-limited settings | Attributed to referral bias or case severity | Conservative fluid management imported from other ALF etiologies compromises the dominant elimination pathway | Poor outcomes following transplant-unit hydration reduction (this trial) |

This table summarizes longstanding clinical paradoxes in amatoxin poisoning and their mechanistic resolution based on findings from this prospective trial. The unifying principle is the kinetic balance between renal clearance and hepatic uptake: under intact glomerular filtration, >80% of systemically circulating amatoxin is rapidly cleared, whereas in oliguric acute kidney injury this fraction redistributes to the liver via arteriohepatic uptake, precipitating multisystem collapse. EHC, enterohepatic circulation; AKI, acute kidney injury; ATN, acute tubular necrosis; ALF, acute liver failure; MELD, Model for End-Stage Liver Disease.

