## Supplementary Appendix for "Elimination Capacity Determines Outcome in Amatoxin Mushroom Poisoning-Induced Acute Liver Failure: A Globally Applicable Management Framework"

**Treatment Guidelines for Amatoxin Mushroom Poisoning**
**The Santa Cruz Protocol**

1) Vomiting and diarrhea beginning 6 hours or more following the ingestion of foraged mushrooms must be considered amatoxin poisoning until proven otherwise. ED treatment and hospital admission should not be delayed pending mycological identification or laboratory confirmation.

2) Labs (CBC, PT/INR, CMP, Lactate) should be repeated every 6 hours. Rapidly rising transaminases (AST, ALT, LDH) confirm the diagnosis. Hemoconcentration, elevated BUN/Creatinine values/ratio, and serum Lactate elevation are typical at presentation. *AMATOXtest* can rapidly confirm the diagnosis from a urine sample.

3) Poison Control Centers should focus the ED solely on the essential task of rapid multi-liter volume replacement and re-establishing urine output. Profuse vomiting and cholera-like diarrhea produces enormous, easily underestimated fluid losses. Aggressive volume replacement reverses hypovolemia induced prerenal-AKI, restores urine output, prevents oliguric-AKI, and slows progression to ALF. Single/multidose activated charcoal, NAC, oral silibinin, Penicillin G Cyclosporine (nephrotoxic) and extracorporeal methods are ineffective interventions that distract precious ED time & resources. Early oliguric AKI development is invariably associated with poor outcomes but easily avoided with sustained aggressive IV hydration.

4) Fluid Management: Place 2 large-bore IVs or multilumen central line & Foley catheter. Bolus LR or NS (4-7 liters) to quickly reestablish brisk urine output and correct vital signs. Then convert to D5 0.45% NS with KCL at 200+ ml/hour (2-3x maintenance).
**Sustained aggressive maintenance IV hydration** dilutes the glomerular filtrate, minimizes renal transit time, enhances elimination, and prevents toxic nephropathy (ATN-induced oliguric-AKI). BUN/Creatinine should correct to values of <10 & <1.0.
Two primary goals:
   a. Maintain brisk urine output (***≥150+ ml/hour*** or 2-3 ml/kg/hour).
   b. Achieve and maintain *serum* ***Lactate*** *correction* (<2).

5) Suspend enterohepatic circulation by maintaining strict NPO (nothing by mouth) status and Octreotide infusion (200 mcg bolus, followed by 50 mcg/hour maintenance). Octreotide tightly seals the Sphincter of Oddi, inhibits gallbladder contraction, and prevents release of amatoxin laden bile into the gut. Discontinue Octreotide and feed the patient once INR recovery is clearly underway, generally ≤30 hours after initiation of SIL infusion or gallbladder aspiration.

6) Serial Lactate values (Lactic acid) are the earliest and most sensitive indicator of prognosis; the INR is more specific. Base clinical decisions on hourly urine outputs, serial Lactates & INR values. The failure of rapid volume replacement and aggressive IV hydration to correct presentation Lactate elevation rarely occurs but is the earliest indication that treatment with SIL will be unsuccessful. A subsequent Lactate rise above 2.2 and/or urine output <150 ml/hour should trigger a bolus of LR or NS and an increase in the maintenance IV fluid rate.

7) Gallbladder Drainage: Performing within 12-18 hours of presentation is ideal; otherwise as early as possible. The gallbladder is the repository of uneliminated amatoxin accumulation. Substantial amatoxin content can still be recovered several days after ingestion. Evacuation of gallbladder bile definitively eliminates amatoxin from the enterohepatic circuit, allowing reversal and clinical recovery from acute liver failure. If SIL cannot be readily obtained, the patient is oliguric, or serum Lactate values remain uncorrected, undertake (by IR if available) Ultrasound-Guided ***Simple Gallbladder Aspiration*** (18 or 19-gauge spinal needle) asap after stabilizing the patient. Risks of perforation, bile peritonitis, bleeding, and infection are substantially reduced compared to ***Percutaneous Cholecystostomy*** drain placement (2$^{nd}$ best alternative). Transhepatic approach if INR <2, otherwise transperitoneal. Consider repeating *Ultrasound-Guided Simple Gallbladder Aspiration* once, 24-48 hours later. *Percutaneous Cholecystostomy* requires leaving a drain in place for several weeks until the tract matures. *Percutaneous Cholecystostomy* is more reliably successful than the third alternative, ***Nasobiliary Drain Placement Under Suction by ERCP*** Must warn the GI endoscopist beforehand not to cut a *sphincterotomy* (hemorrhage risk). Provide ***FFP*** before these procedures if the INR is elevated (≥1.7). ***Surgical cholecystostomy*** can be performed if the other procedures are unavailable or fail to recover gallbladder bile. Amatoxin-laden bile looks like burnt motor oil. Freeze recovered bile for future diagnostic analysis.

***Preferred Technique*** (see Figure 1)

***GB aspiration using Cook 21g needle and 5 Fr Micropuncture Introducer Set** under ultrasound & fluoroscopy. First aspirate and collect the bile; then inject 10 ml NS and repeat the process until the aspirate is mostly clear. Remove the catheter. Optionally, place Gel Foam to close the tract through the sheath (see photos below). Can be repeated after 24-48 hours if necessary.

8) SIL (Silibinin dihemisuccinate, aka Legalon SIL), 5 mg/kg bolus, followed by 20 mg/kg/day continuous infusion, produces INR reduction by ~infusion hour 36, reliably heralding complete clinical recovery, assuming sustained serum Lactate correction and good urine output. Infuse 24-96 hours. Hospital LOS averages 5-6 days. Warmth or flushing during the initial bolus is the only commonly observed adverse effect. The SIL initiation window is ~120 hours post-ingestion, assuming preservation of renal function and sustained serum lactate correction. SIL is no longer available under Open IND. To obtain SIL emergently (USA) call FDA Office of Emergency Operations: (301)-796-8240. Shipping from Viatris (East Coast) takes ~24-36 hours.

9) **Hepatic Encephalopathy**: Lactulose administered rectally is the treatment of choice. If CRRT is considered for this indication (*HE*) care must be taken to ensure no compromise in the rate of IV fluid infusion or urine output. Interruption, suspension, or significant reduction in IV hydration markedly increases the risk of developing ***ATN-related oliguric-AKI*** and a poor outcome.

**S Todd Mitchell MD, MPH**
**Amatoxin Mushroom Poisoning**

+1-(831)-588-2192

Figure 1. Gallbladder Aspiration and Percutaneous Cholecystostomy by Interventional Radiology under ultrasound and fluoroscopy guidance.

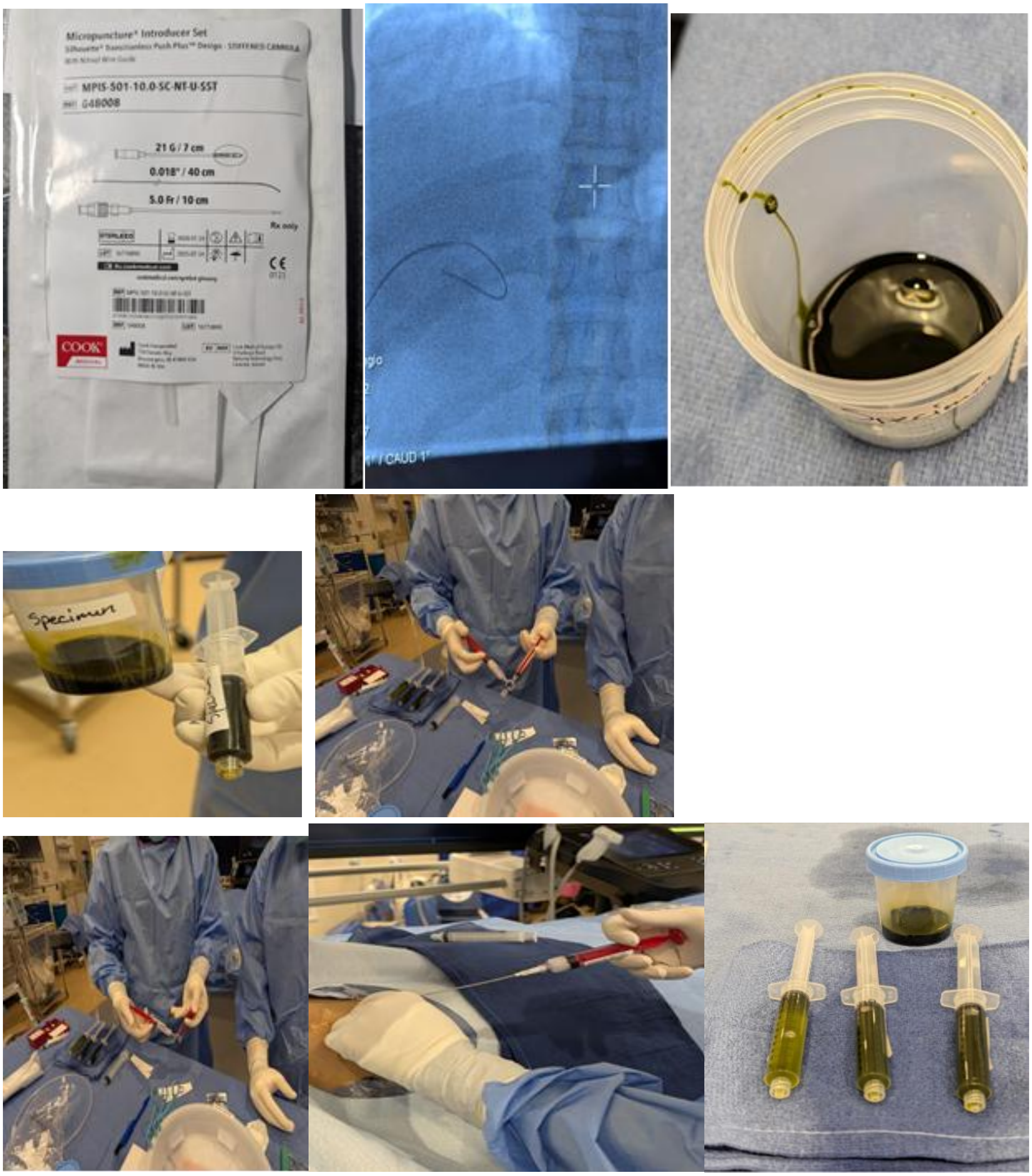

### Listing S-1. All Patients Studied, Safety Population (n=99)

| | | | Hours from Ingestion to | | | SIL | | | Additional Treaatment Used | | | | LOS | Patient | Maximum observed laboratory value | | | | | | | | | | Population | |
|---|---|---|---|---|---|---|---|---|---|---|---|---|---|---|---|---|---|---|---|---|---|---|---|---|---|---|
| Patient ID | Age/Sex | Race | Symptoms | First lab | Start SIL | Dose (mg/kg) | Duration (days) | Hydration Restricted | NAC | Activated Charcoal | Octreotide | Bile drainage | Hospital days | Outcome | Hgb | BUN | Cr | AST | ALT | Total Bilirubin | INR | Lactate | MELD Score | Protocol version | IND | Efficacy |
| 180 | 10-14/M | Caucasian | 10.5 | 42.5 | 66.5 | 25.0 | 1.0 | 0 | 1 | 1 | 0 | 0 | 6.0 | Discharge | | | | 194 | 320 | | 1 | | | 0 | 0 | 1 |
| 181 | 10-14/M | Caucasian | 10.5 | 42.5 | 66.5 | 25.0 | 1.0 | 0 | 1 | 1 | 0 | 0 | 6.0 | Discharge | | | | 33 | 197 | | 1 | | | 0 | 0 | 1 |
| 182 | 80-84/F | Caucasian | 10.5 | 30.6 | 66.5 | 5.0 | 0.1 | 0 | 1 | 1 | 0 | 0 | 6.0 | Discharge | 14 | 25 | 0.93 | 3184 | 4873 | 1.2 | 1.6 | | 12.4 | 0 | 0 | 0 |
| 183 | 55-59/M | Caucasian | 16.0 | 71.3 | 66.0 | 45.0 | 2.0 | 0 | 1 | 0 | 0 | 0 | 4.0 | Discharge | | 19 | 0.95 | 3213 | 6101 | 1 | 1.6 | | 11.7 | 0 | 0 | 1 |
| 184 | 25-29/M | Hispanic | 11.0 | 15.5 | 75.0 | 45.0 | 2.0 | 0 | 1 | 1 | 0 | 0 | 6.0 | Discharge | 16 | 16 | 1.2 | 332 | 554 | 1.7 | 1.3 | | 13.1 | 0 | 0 | 1 |
| 185 | 15-19/F | Hispanic | 8.0 | 15.8 | 73.3 | 45.0 | 2.0 | 0 | 1 | 1 | 0 | 0 | 6.0 | Discharge | 13 | 17 | 0.8 | 2196 | 3895 | 6.6 | 2.2 | | 22.4 | 0 | 0 | 1 |
| 186 | 65-69/M | Hispanic | 11.0 | 13.5 | 74.6 | 45.0 | 2.0 | 0 | 1 | 1 | 0 | 0 | 6.0 | Discharge | | | | 6790 | 8553 | 7.9 | 3.6 | | | 0 | 0 | 1 |
| 187 | 55-59/F | Hispanic | 11.0 | 13.5 | 75.0 | 45.0 | 2.0 | 0 | 1 | 1 | 0 | 0 | 6.0 | Discharge | 16 | 34 | 1.2 | 6790 | 9476 | 7.9 | 3.8 | | 30.9 | 0 | 0 | 1 |
| 188 | 85-89/F | Hispanic | 11.0 | 14.3 | 75.0 | 85.0 | 4.0 | 1 | 1 | 1 | 0 | 0 | 10.0 | Death | 13 | 30 | 5.6 | 19972 | 12224 | 13.8 | 5.9 | | 49.5 | 0 | 0 | 0 |
| 189 | 30-34/M | Hispanic | 8.0 | 15.7 | 75.0 | 125.0 | 6.0 | 0 | 1 | 1 | 0 | 0 | 11.0 | Discharge | 17 | 16 | 1.1 | 15104 | 18073 | 19.6 | 14.9 | | 48.8 | 0 | 0 | 1 |
| 1 | 30-34/F | Hispanic | 9.0 | 53.5 | 63.2 | 120.8 | 5.1 | 1 | 1 | 1 | 0 | 0 | 13.0 | Death | 17 | 22 | 3.7 | 4350 | 3995 | 19.9 | 3.4 | 10.7 | 44.0 | 1 | 1 | 0 |
| 2 | 35-39/F | Hispanic | 9.0 | 53.7 | 63.1 | 122.4 | 5.1 | 1 | 1 | 1 | 0 | 0 | 13.0 | LT | 12 | 4 | 2 | 10247 | 6634 | 11.7 | 8.9 | 5.7 | 46.8 | 1 | 1 | 0 |
| 3 | 35-39/M | Hispanic | 9.0 | 50.4 | 56.0 | 120.0 | 5.4 | 1 | 1 | 1 | 0 | 1 | 13.0 | Discharge | 17 | 22 | 2.7 | 11590 | 7151 | 17 | 4.6 | 6.1 | 43.7 | 1 | 1 | 0 |
| 4 | 45-49/M | Caucasian | 9.0 | 17.5 | 49.0 | 98.1 | 4.0 | 0 | 1 | 1 | 0 | 0 | 7.0 | Discharge | 15 | 21 | 0.9 | 2204 | 3854 | 2 | 1.3 | 3.5 | 12.0 | 1 | 1 | 1 |
| 5 | 70-74/F | Other | 14.0 | 35.3 | 63.0 | 112.9 | 5.0 | 0 | 1 | 1 | 0 | 0 | 9.0 | Discharge | 13 | 20 | 1.1 | 8658 | 8160 | 8.7 | 3.3 | | 28.9 | 1 | 1 | 1 |
| 6 | 75-79/M | Caucasian | 15.0 | 23.8 | 63.8 | 66.7 | 3.4 | 0 | 0 | 1 | 0 | 1 | 7.0 | Discharge | 16 | 29 | 1.4 | 1382 | 2502 | 6 | 1.3 | | 19.4 | 1 | 1 | 1 |
| 7 | 45-49/M | Other | 14.0 | 29.5 | 49.4 | 73.4 | 2.8 | 0 | 1 | 1 | 0 | 0 | 6.0 | Discharge | 17 | 27 | 0.9 | 1575 | 2728 | 1.34 | 1.4 | | 11.3 | 1.1 | 1 | 1 |
| 8 | 45-49/M | Chinese | 11.0 | 50.0 | 50.0 | 75.0 | 3.0 | 0 | 1 | 1 | 0 | 0 | 7.0 | Discharge | 15 | 6 | 0.7 | 4830 | 5570 | 3.32 | 1.32 | | 14.1 | 1.1 | 1 | 1 |
| 9 | 70-74/F | Other | 12.0 | 73.0 | 98.5 | 25.1 | 1.0 | 0 | 1 | 1 | 0 | 0 | 5.0 | Discharge | 13 | 20 | 0.8 | 8726 | 7210 | 5.1 | 3 | | 24.9 | 1.1 | 1 | 1 |
| 10 | 40-44/F | Other | 14.0 | 18.8 | 98.3 | 24.6 | 1.0 | 0 | 1 | 0 | 0 | 0 | 5.0 | Discharge | 16 | 19 | 0.7 | 4449 | 5112 | 1.5 | 1.7 | 1.9 | 13.9 | 1.1 | 1 | 1 |
| 11 | 40-44/F | Caucasian | 10.5 | 37.2 | 54.0 | 181.7 | 8.5 | 0 | 1 | 0 | 0 | 0 | 15.0 | Discharge | 17 | 64 | 8.83 | 18117 | 8966 | 9.1 | 2.4 | 2.7 | 37.8 | 1.1 | 1 | 1 |
| 12 | 75-79/F | Other | 12.0 | 22.6 | 55.8 | 46.6 | 1.5 | 0 | 1 | 1 | 0 | 0 | 6.0 | Discharge | 14 | 27 | 0.98 | 32 | 35 | 0.9 | 1.2 | | 8.5 | 1.1 | 1 | 1 |
| 13 | 80-84/M | Other | 12.0 | 22.3 | 55.7 | 46.6 | 2.5 | 0 | 1 | 1 | 0 | 0 | 6.0 | Discharge | 14 | 32 | 1.73 | 207 | 198 | 1.9 | 1.5 | | 18.6 | 1.1 | 1 | 1 |
| 14 | 60-64/M | Caucasian | 10.0 | 22.5 | 51.3 | 61.8 | 2.4 | 0 | 1 | 1 | 0 | 0 | 6.0 | Discharge | 18 | 26 | 2.19 | 114 | 55 | 0.6 | 1.1 | | 15.0 | 1.1 | 1 | 1 |
| 15 | 45-49/F | Other | 12.0 | 80.3 | 107.1 | 66.5 | 1.6 | 0 | 1 | 1 | 0 | 0 | 6.0 | Discharge | 12 | 7 | 0.75 | 2488 | 5649 | 1.8 | 1.2 | | 10.7 | 1.1 | 1 | 1 |
| 16 | 70-74/F | Chinese | 12.0 | 37.0 | 40.0 | 30.6 | 2.0 | 0 | 0 | 0 | 0 | 0 | 6.0 | Discharge | 14 | 10 | 0.72 | 258 | 266 | 1.2 | 1.1 | | 8.2 | 1.1 | 1 | 1 |
| 17 | 70-74/M | Chinese | 14.0 | 26.8 | 41.0 | 79.6 | 4.0 | 0 | 0 | 1 | 0 | 0 | 7.0 | Discharge | 17 | 24 | 1.13 | 7098 | 6778 | 8 | 2.1 | | 23.8 | 1.1 | 1 | 1 |
| 18 | 5-9/M | Chinese | 25.5 | 55.5 | 61.7 | 105.0 | 4.7 | 0 | 1 | 0 | 0 | 0 | 8.0 | Discharge | 13 | 11 | 0.31 | 12448 | 11476 | 3.1 | 4.7 | | 28.0 | 1.1 | 1 | 1 |
| 19 | 50-54/M | Caucasian | 6.5 | 15.5 | 19.0 | 62.9 | 3.3 | 0 | 0 | 0 | 0 | 1 | 5.0 | Discharge | 17 | 23 | 0.9 | 6569 | 7953 | 6.6 | 1.8 | | 20.1 | 1.1 | 1 | 1 |
| 20 | 50-54/M | Caucasian | 11.5 | 61.2 | 70.5 | 27.7 | 1.7 | 0 | 0 | 0 | 0 | 0 | 6.0 | Discharge | 15 | 21 | 0.7 | 4387 | 8561 | 3.5 | 2 | | 18.9 | 1.1 | 1 | 1 |
| 21 | 65-69/M | Caucasian | 12.0 | 46.3 | 123.0 | 61.2 | 2.8 | 0 | 0 | 0 | 0 | 0 | 17.0 | LT | 19 | 41 | 2.3 | 4442 | 7870 | 15.3 | 5.8 | 6.6 | 44.4 | 1.1 | 1 | 0 |
| 22 | 50-54/M | Caucasian | 17.0 | 55.9 | 65.0 | 66.7 | 3.0 | 0 | 0 | 0 | 0 | 0 | 6.0 | Discharge | 17 | 13 | 0.81 | 10711 | 27117 | 7.9 | 1.7 | | 20.2 | 1.1 | 1 | 1 |
| 23 | 55-59/M | Caucasian | 17.0 | 49.3 | 73.3 | 25.2 | 1.0 | 0 | 0 | 0 | 0 | 0 | 4.0 | Discharge | 16 | 30 | 1.03 | 485 | 613 | 0.8 | 1 | | 6.7 | 1.1 | 1 | 1 |
| 24 | 40-44/F | Other | 24.0 | 128.3 | 146.0 | 54.9 | 2.9 | 0 | 1 | 0 | 0 | 0 | 5.0 | Death | 14 | 15 | 3.7 | 5420 | 5658 | 13.1 | 14.1 | 12.1 | 58.3 | 1.1 | 1 | 0 |
| 25 | 75-79/F | Other | 7.5 | 28.8 | 30.0 | 75.8 | 2.6 | 0 | 1 | 1 | 0 | 0 | 5.0 | Discharge | 15 | 12 | 0.51 | 5403 | 5297 | 7.5 | 4 | | 29.6 | 1.1 | 1 | 1 |
| 26 | 50-54/M | Other | 14.5 | 73.6 | 85.5 | 40.0 | 1.1 | 0 | 1 | 1 | 0 | 1 | 5.0 | Discharge | 14 | 43 | 1.88 | 1258 | 4400 | 1.7 | 1.3 | 1 | 17.4 | 1.1 | 1 | 1 |
| 27 | 80-84/M | Caucasian | 10.0 | 60.0 | 61.0 | 117.5 | 5.5 | 0 | 1 | 1 | 0 | 0 | 7.0 | Discharge | 15 | 44 | 1.58 | 1995 | 3384 | 7.7 | 2.5 | | 28.8 | 1.1 | 1 | 1 |
| 28 | 45-49/F | Other | 12.0 | 73.2 | 77.2 | 79.6 | 2.6 | 0 | 0 | 0 | 0 | 0 | 5.0 | Discharge | 13 | 4 | 0.57 | 3638 | 4405 | 2.5 | 2.3 | | 19.2 | 1.1 | 1 | 1 |
| 29 | 55-59/F | Other | 12.0 | 73.3 | 79.5 | 25.0 | 1.0 | 0 | 0 | 0 | 0 | 0 | 3.0 | Discharge | 17 | 15 | 0.9 | 4018 | 5008 | 1.3 | 1.3 | | 10.4 | 1.1 | 1 | 1 |
| 30 | 70-74/F | Other | 26.0 | 21.8 | 52.7 | 29.9 | 1.2 | 1 | 1 | 1 | 0 | 0 | 3.0 | Death | 14 | 34 | 1.85 | 1275 | 1430 | 2.7 | 6.58 | 7.1 | 37.2 | 1.1 | 1 | 0 |
| 31 | 55-59/F | Other | 14.0 | 60.2 | 73.7 | 30.5 | 1.8 | 0 | 1 | 1 | 0 | 0 | 5.0 | Discharge | 14 | 14 | 0.75 | 4556 | 8681 | 2.3 | 1.69 | | 15.5 | 1.1 | 1 | 1 |
| 32 | 80-84/F | Other | 13.5 | 32.1 | 54.0 | 72.1 | 2.7 | 0 | 1 | 1 | 0 | 0 | 6.0 | Discharge | 14 | 20 | 0.8 | 3291 | 4138 | 3.2 | 3 | | 23.1 | 1.1 | 1 | 1 |
| 33 | 55-59/M | Caucasian | 11.0 | 41.5 | 51.5 | 43.2 | 1.6 | 0 | 0 | 0 | 0 | 0 | 4.0 | Discharge | 19 | 61 | 2.9 | 2481 | 2701 | 2.5 | 1.3 | | 23.0 | 1.1 | 1 | 1 |
| 34 | 55-59/F | Chinese | 12.0 | 41.5 | 54.0 | 221.4 | 10.5 | 0 | 1 | 1 | 0 | 0 | 13.0 | Death | 19 | 54 | 3 | 1709 | 3012 | 25.8 | 14 | 16.86 | 58.8 | 1.1 | 1 | 0 |
| 35 | 5-9/M | Other | 8.0 | 27.5 | 52.0 | 40.6 | 1.1 | 0 | 1 | 1 | 0 | 0 | 4.0 | Discharge | 14 | 29 | 0.93 | 410 | 363 | 0.8 | 1.3 | | 9.4 | 1.1 | 1 | 1 |
| 36 | 70-74/M | Caucasian | 14.0 | 67.5 | 93.0 | 35.4 | 2.9 | 0 | 1 | 1 | 0 | 0 | 5.0 | Discharge | 17 | 39 | 1.7 | 2009 | 3575 | 2.4 | 1.3 | | 17.8 | 1.1 | 1 | 1 |
| 37 | 20-24/M | Caucasian | 24.0 | 125.6 | 149.7 | 19.9 | 1.0 | 0 | 1 | 0 | 0 | 1 | 3.0 | LT | 17 | 30 | 0.87 | 5510 | 6878 | 9 | 14 | 5.5 | 44.3 | 1.1 | 1 | 0 |
| 38 | 25-29/F | Chinese | 14.0 | 22.2 | 41.0 | 85.0 | 4.0 | 0 | 1 | 0 | 0 | 0 | 7.0 | Discharge | 14 | 14 | 0.63 | 114 | 155 | 2.1 | 1.3 | | 12.2 | 1.1 | 1 | 1 |
| 39 | 55-59/F | Chinese | 14.0 | 22.2 | 41.0 | 83.7 | 4.0 | 0 | 1 | 0 | 0 | 0 | 7.0 | Discharge | 15 | 16 | 0.63 | 1312 | 2306 | 2.4 | 1.4 | | 13.5 | 1.1 | 1 | 1 |
| 40 | 90-94/F | Caucasian | 9.0 | 83.0 | 91.8 | 89.8 | 3.9 | 0 | 1 | 1 | 0 | 0 | 10.0 | Discharge | 14 | 18 | 1.4 | 512 | 521 | 3 | 1.6 | | 19.1 | 1.1 | 1 | 1 |

| | | | Hours from Ingestion to | | | SIL | | | Additional Treaatment Used | | | | LOS | Patient | Maximum observed laboratory value | | | | | | | | | | Population | |
|---|---|---|---|---|---|---|---|---|---|---|---|---|---|---|---|---|---|---|---|---|---|---|---|---|---|---|
| Patient ID | Age/Sex | Race | Symptoms | First lab | Start SIL | Dose (mg/kg) | Duration (days) | Hydration Restricted | NAC | Activated Charcoal | Octreotide | Bile drainage | Hospital days | Outcome | Hgb | BUN | Cr | AST | ALT | Total Bilirubin | INR | Lactate | MELD Score | Protocol version | IND | Efficacy |
| 41 | 60-64/M | Caucasian | 8.0 | 28.1 | 40.0 | 125.0 | 5.0 | 0 | 1 | 1 | 0 | 0 | 16.0 | Discharge | 21 | 62 | 5.39 | 1401 | 2425 | 19.3 | 2.8 | | 42.4 | 1.1 | 1 | 1 |
| 42 | 55-59/M | Caucasian | 6.0 | 26.4 | 40.0 | 25.0 | 1.0 | 0 | 1 | 1 | 0 | 0 | 5.0 | Discharge | 16 | 16 | 0.81 | 173 | 234 | 0.9 | 1.2 | | 8.5 | 1.1 | 1 | 1 |
| 43 | 45-49/M | Caucasian | 6.0 | 28.8 | 40.0 | 25.0 | 1.0 | 0 | 1 | 1 | 0 | 0 | 9.0 | Discharge | 17 | 23 | 0.92 | 101 | 103 | 0.8 | 1.1 | | 7.5 | 1.1 | 1 | 1 |
| 44 | 60-64/M | Other | 12.0 | 64.0 | 88.0 | 42.0 | 1.5 | 0 | 1 | 1 | 0 | 0 | 3.0 | Discharge | 21 | 61 | 4.2 | 351 | 1332 | 2 | 1.2 | 3.83 | 24.4 | 1.1 | 1 | 1 |
| 45 | 60-64/M | Caucasian | 12.0 | 71.9 | 88.0 | 74.0 | 3.0 | 0 | 1 | 1 | 0 | 0 | 11.0 | Discharge | 14 | 39 | 1.39 | 12137 | 5194 | 10.1 | 2.8 | | 29.9 | 1.1 | 1 | 1 |
| 46 | 25-29/F | Other | 11.0 | 20.1 | 73.0 | 60.0 | 1.6 | 0 | 1 | 1 | 0 | 0 | 7.0 | Discharge | 14 | 12 | 0.7 | 1554 | 2049 | 1.1 | 1.5 | | 11.3 | 1.1 | 1 | 1 |
| 47 | 60-64/F | Chinese | 10.0 | 44.8 | 55.0 | 81.8 | 2.5 | 1 | 1 | 1 | 0 | 0 | 5.0 | Death | 17 | 48 | 2.8 | 8160 | 6809 | 3.2 | 13.8 | 26 | 50.1 | 1.1 | 1 | 0 |
| 48 | 80-84/F | Other | 12.0 | 17.6 | 49.0 | 53.0 | 1.8 | 0 | 1 | 1 | 0 | 0 | 4.0 | Discharge | 16 | 16 | 0.9 | 77 | 122 | 1.7 | 1.2 | | 10.5 | 1.1 | 1 | 1 |
| 49 | 95-99/F | Caucasian | 24.0 | 69.5 | 82.5 | 33.5 | 2.6 | 0 | 1 | 0 | 0 | 0 | 6.0 | Discharge | 13 | 21 | 1.01 | 3141 | 2633 | 2.8 | 2.6 | | 21.1 | 1.1 | 1 | 1 |
| 50 | 40-44/F | Chinese | 10.0 | 26.3 | 52.0 | 85.0 | 2.6 | 0 | 0 | 0 | 0 | 1 | 5.0 | Discharge | 13 | 21 | 1.1 | 3827 | 6173 | 3 | 2.5 | | 21.8 | 1.1 | 1 | 1 |
| 51 | 65-69/M | Chinese | 14.0 | 23.1 | 40.4 | 74.6 | 2.4 | 0 | 1 | 1 | 1 | 0 | 8.0 | Discharge | 14 | 18 | 0.98 | 5102 | 3475 | 8.2 | 1.9 | | 21.6 | 1.2 | 1 | 1 |
| 52 | 50-54/F | Chinese | 13.5 | 47.8 | 65.5 | 82.1 | 3.0 | 0 | 1 | 1 | 1 | 0 | 8.0 | Discharge | 12 | 5 | 0.62 | 8000 | 8270 | 6.8 | 6.3 | 3.4 | 34.3 | 1.2 | 1 | 1 |
| 53 | 80-84/M | Chinese | 19.0 | 53.8 | 65.5 | 60.2 | 2.0 | 0 | 1 | 0 | 1 | 0 | 6.0 | Discharge | 12 | 9 | 0.74 | 2630 | 3270 | 6.1 | 2.8 | 3.9 | 24.8 | 1.2 | 1 | 1 |
| 54 | 75-79/F | Chinese | 13.5 | 52.1 | 65.5 | 40.3 | 1.5 | 0 | 1 | 0 | 1 | 0 | 5.0 | Discharge | 13 | 9 | 0.64 | 413 | 616 | 1.4 | 1.5 | 2.7 | 12.2 | 1.2 | 1 | 1 |
| 55 | 25-29/F | Chinese | 9.0 | 28.2 | 51.0 | 104.5 | 3.8 | 0 | 1 | 1 | 1 | 0 | 6.0 | Discharge | 14 | 19 | 0.75 | 3274 | 5596 | 5.2 | 3.1 | | 25.3 | 1.2 | 1 | 1 |
| 56 | 50-54/M | Chinese | 13.0 | 31.0 | 51.0 | 85.8 | 2.8 | 0 | 1 | 1 | 1 | 0 | 6.0 | Discharge | 15 | 21 | 0.92 | 2535 | 2968 | 1.7 | 1.9 | | 15.6 | 1.2 | 1 | 1 |
| 57 | 70-74/M | Chinese | | 61.0 | 63.6 | 70.9 | 0.9 | 0 | 1 | 1 | 1 | 0 | 4.0 | Discharge | 19 | 21 | 1 | 2314 | 3983 | 4.8 | 1.8 | 1.9 | 18.9 | 1.2 | 1 | 1 |
| 58 | 70-74/M | Other | 14.0 | 54.4 | 76.2 | 64.2 | 1.8 | 0 | 1 | 1 | 1 | 0 | 6.0 | Discharge | 12 | 25 | 1.3 | 6427 | 6163 | 4.3 | 2.7 | 5 | 25.6 | 1.2 | 1 | 1 |
| 59 | 65-69/F | Chinese | 11.0 | 16.8 | 42.0 | 26.1 | 1.0 | 0 | 1 | 1 | 1 | 0 | 12.0 | Discharge | 16 | 17 | 0.7 | 4122.1 | 3740.1 | 4.9 | 4.3 | 4 | 28.8 | 1.2 | 1 | 1 |
| 60 | 55-59/F | Chinese | 18.0 | 22.5 | 42.0 | 45.0 | 2.1 | 0 | 1 | 1 | 1 | 0 | 4.0 | Discharge | 14 | 17 | 0.53 | 15 | 31 | 0.9 | 1.1 | 3.3 | 7.5 | 1.2 | 1 | 1 |
| 61 | 55-59/M | Chinese | 14.5 | 28.5 | 44.4 | 170.0 | 3.1 | 0 | 1 | 1 | 1 | 0 | 6.0 | Discharge | 15 | 13 | 0.97 | 6195 | 5505 | 13 | 3.2 | 3.1 | 29.2 | 1.2 | 1 | 1 |
| 62 | 70-74/F | Caucasian | 17.0 | 45.0 | 56.0 | 60.4 | 2.5 | 0 | 0 | 0 | 1 | 0 | 4.0 | Discharge | 13 | 36 | 2.3 | 60 | 80 | 1.3 | 1.1 | 1.4 | 16.5 | 1.2 | 1 | 1 |
| 63 | 85-89/M | Caucasian | 8.5 | 42.9 | 55.9 | 64.8 | 2.5 | 0 | 0 | 0 | 1 | 0 | 6.0 | Discharge | 13 | 45 | 1.8 | 1242 | 3305 | 2.1 | 2.2 | 1.6 | 23.7 | 1.2 | 1 | 1 |
| 64 | 45-49/M | Other | 6.8 | 45.5 | 65.4 | 61.1 | 2.4 | 0 | 0 | 0 | 1 | 0 | 5.0 | Discharge | 14 | 10 | 0.78 | 1948 | 2303 | 2.7 | 1.6 | 1.74 | 15.4 | 1.3 | 1 | 1 |
| 65 | 65-69/M | Other | 24.0 | 58.5 | 75.0 | 105.0 | 4.9 | 0 | 1 | 1 | 1 | 0 | 7.0 | Discharge | 15 | 11 | 0.85 | 3601 | 4653 | 2.6 | 1.9 | 1.7 | 17.2 | 1.3 | 1 | 1 |
| 66 | 60-64/F | Caucasian | 12.0 | 17.8 | 61.2 | 53.6 | 2.2 | 0 | 1 | 1 | 1 | 0 | 4.0 | Discharge | 15 | 19 | 0.7 | 2155 | 3569 | 1.2 | 1.9 | 3 | 14.3 | 1.3 | 1 | 1 |
| 67 | 60-64/F | Chinese | 12.0 | 50.0 | 74.5 | 85.0 | 3.0 | 0 | 1 | 0 | 1 | 0 | 6.0 | Discharge | 15 | 35 | 0.97 | 3353 | 4505 | 3.1 | 3.2 | 2.6 | 23.7 | 1.3 | 1 | 1 |
| 68 | 75-79/M | Caucasian | 11.0 | 25.3 | 72.0 | 65.0 | 3.0 | 0 | 1 | 0 | 1 | 0 | 7.0 | Discharge | 14 | 26 | 1 | 3236 | 5590 | 4.9 | 2 | 2.3 | 20.2 | 1.3 | 1 | 1 |
| 69 | 45-49/F | Chinese | 15.0 | 52.7 | 80.0 | 25.0 | 1.6 | 0 | 0 | 0 | 1 | 0 | 6.0 | Discharge | 13 | 7 | 0.68 | 7700 | 7080 | 5.7 | 2.29 | 2.3 | 22.3 | 1.3 | 1 | 1 |
| 70 | 10-14/M | Other | 14.0 | 57.0 | 78.0 | 55.4 | 2.5 | 0 | 1 | 0 | 1 | 0 | 11.0 | Discharge | 12 | 9 | 0.48 | 12232 | 7560 | 5 | 3.8 | 5.2 | 27.5 | 1.3 | 1 | 1 |
| 71 | 55-59/M | Other | 14.0 | 56.7 | 101.0 | 25.0 | 0.0 | 0 | 1 | 0 | 1 | 0 | 3.0 | Discharge | 17 | 42 | 4.79 | 1551 | 1103 | 2.3 | 1.2 | 2.1 | 24.9 | 1.3 | 1 | 1 |
| 72 | 55-59/M | Caucasian | | 45.0 | 93.0 | 42.4 | 2.5 | 0 | 1 | 0 | 1 | 0 | 6.0 | Discharge | 18 | 11 | 1.67 | 4011 | 3097 | 2.2 | 1.7 | 3 | 20.3 | 1.3 | 1 | 1 |
| 73 | 20-24/M | Caucasian | 24.0 | 71.6 | 103.5 | 75.3 | 4.0 | 0 | 1 | 0 | 1 | 0 | 5.5 | Discharge | 16 | 55 | 2.92 | 4196 | 6815 | 13.6 | 5.8 | 3.8 | 46.2 | 1.3 | 1 | 1 |
| 74 | 50-54/M | Chinese | 12.0 | 15.9 | 118.3 | 65.1 | 5.4 | 0 | 1 | 1 | 1 | 0 | 10.5 | Discharge | 17 | 30 | 1.84 | 6777 | 8161 | 17.6 | 6.1 | 6.7 | 43.4 | 1.3 | 1 | 1 |
| 75 | 35-39/M | Hispanic | 12.0 | 19.3 | 45.5 | 85.0 | 4.0 | 0 | 1 | 0 | 1 | 0 | 7.0 | Discharge | 19 | 27 | 1.4 | 3084 | 6999 | 5.1 | 3.2 | 2.6 | 28.8 | 1.3 | 1 | 1 |
| 76 | 0-4/F | Other | 12.0 | 20.0 | 15.0 | 49.9 | 3.7 | 1 | 0 | 0 | 1 | 0 | 38.0 | LT | 11 | 16 | 0.4 | 14300 | 10200 | 11.4 | 10.2 | 10.4 | 41.6 | 1.3 | 1 | 0 |
| 77 | 25-29/F | Other | 10.0 | 19.8 | 40.9 | 107.1 | 5.2 | 0 | 0 | 0 | 1 | 0 | 6.0 | Discharge | 17 | 21 | 1.01 | 11427 | 9833 | 5.4 | 3.2 | 5.87 | 25.9 | 1.3 | 1 | 1 |
| 78 | 30-34/M | Other | 10.0 | 23.2 | 37.8 | 50.3 | 2.3 | 0 | 0 | 0 | 1 | 0 | 14.0 | Discharge | 17 | 22 | 1.46 | 6123 | 4703 | 3.3 | 1.4 | 3.5 | 18.3 | 1.3 | 1 | 1 |
| 79 | 40-44/F | Other | 12.0 | 36.1 | 39.3 | 42.3 | 1.9 | 0 | 0 | 0 | 1 | 1 | | LT | 16 | 24 | 0.8 | 9573 | 6239 | 5.6 | 7.8 | 7.4 | 35.9 | 1.3 | 1 | 1 |
| 80 | 50-54/F | Other | 12.0 | 48.8 | 71.1 | 99.8 | 4.8 | 0 | 0 | 0 | 1 | 0 | 5.0 | Discharge | 14 | 52 | 2.24 | 11940 | 11350 | 13.6 | 4.5 | 8 | 40.9 | 1.3 | 1 | 1 |
| 81 | 35-39/M | Caucasian | 7.5 | 11.7 | 28.3 | 45.0 | 2.0 | 0 | 0 | 0 | 1 | 0 | 6.0 | Discharge | 17 | 18 | 1.3 | 1948 | 3851 | 2.4 | 1.6 | 2.2 | 17.5 | 1.3 | 1 | 1 |
| 82 | 55-59/M | Hispanic | 12.0 | 61.7 | 78.5 | 15.0 | 0.5 | 1 | 1 | 0 | 1 | 0 | 2.2 | LT | 21 | 61 | 2.33 | 2542 | 8145 | 6.9 | 9.01 | 6.8 | 46.4 | 1.3 | 1 | 0 |
| 83 | 75-79/M | Other | 11.5 | 21.9 | 37.5 | 45.0 | 2.0 | 0 | 1 | 0 | 1 | 0 | 4.0 | Discharge | 17 | 22 | 2.3 | 288 | 397 | 0.7 | 1.6 | 1.9 | 19.7 | 1.3 | 1 | 1 |
| 84 | 45-49/F | Other | 8.0 | 12.5 | 31.8 | 87.8 | 4.2 | 0 | 1 | 1 | 1 | 0 | 5.0 | Discharge | 14 | 15 | 0.7 | 3840 | 5724 | 4.4 | 1.9 | 2.9 | 19.2 | 1.3 | 1 | 1 |
| 85 | 60-64/M | Other | 16.0 | 31.3 | 62.0 | 48.4 | 2.2 | 0 | 1 | 1 | 1 | 0 | 4.0 | Discharge | 16 | 27 | 1.8 | 2847 | 2145 | 2.7 | 1.9 | 2.8 | 23.0 | 1.3 | 1 | 1 |
| 86 | 25-29/M | Caucasian | 24.0 | 61.2 | 95.5 | 55.0 | 2.5 | 0 | 1 | 0 | 1 | 0 | 6.0 | Discharge | 14 | 35 | 6.42 | 12288 | 7378 | 17.4 | 6 | 6.2 | 50.6 | 1.3 | 1 | 1 |
| 87 | 75-79/M | Chinese | 7.0 | 30.0 | 77.0 | 45.0 | 2.0 | 0 | 1 | 0 | 1 | 0 | 6.0 | Discharge | 18 | 38 | 1.8 | 4876 | 5551 | 6.1 | 2.7 | 5.3 | 30.0 | 1.3 | 1 | 1 |
| 291 | 85-89/M | Caucasian | 12.0 | 31.7 | 69.0 | 62.6 | 2.7 | 0 | 1 | 1 | 0 | 0 | 11.0 | Discharge | 16 | 40 | 2.5 | 3544 | 3501 | 3.7 | 1.9 | | 27.3 | 1.1 | 0 | 1 |
| 292 | 80-84/M | Caucasian | 17.0 | 59.7 | 91.0 | 85.0 | 2.9 | 0 | 1 | 1 | 1 | 0 | 8.0 | Discharge | 17 | 43.42 | 1.39 | 1559 | 3163 | 7.7778 | 2 | | 25.1 | 1.2 | 0 | 1 |

SIL = IV silibinin, NAC = n-acetyl cysteine, LOS = length of stay, LT = liver transplant, MELD (Model for End-Stage Liver Disease) score=3.78·ln(Bili)+11.2·ln(INR)+9.57·ln(Cr)+6.43; Protocol version: 0=emergency IND, 1=original protocol, 1.1=added OSI commitment form, 1.2=added octreotide & lactate, 1.3=added treatment guidelines requirement

**Figure S-1. Transplant-Free Recovery vs. MELD Score**
Logistic Fit of Outcome by MELD score

#### Safety Population (n=99)

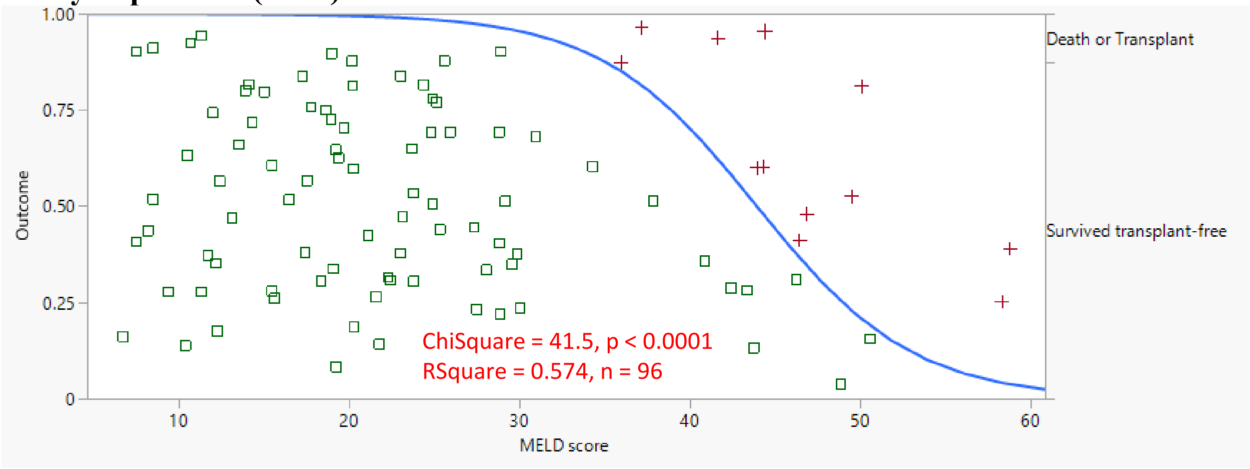

#### Efficacy Population (n=86)

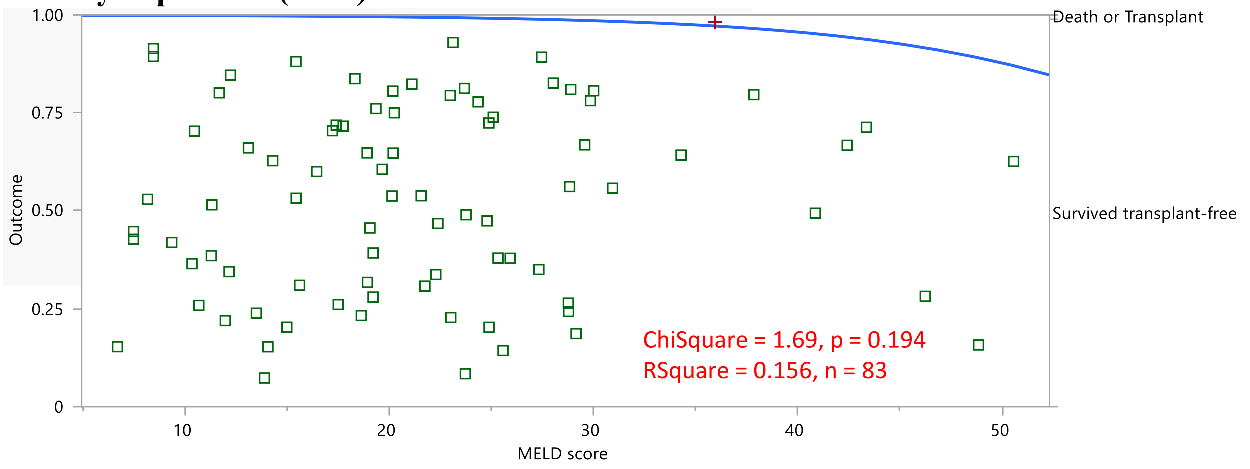

MELD (Model for End-Stage Liver Disease) score = 3.78·ln(Bili)+11.2·ln(INR)+9.57·ln(Cr)+6.43 is often capped at 40 for prediction of survival, shown here without the 40 cap.

**Figure S-2. Survival Analysis for Death or Transplant**

For the 12 patients with an outcome of death or liver transplant

median [95% CI] 179 [115, 281] hours

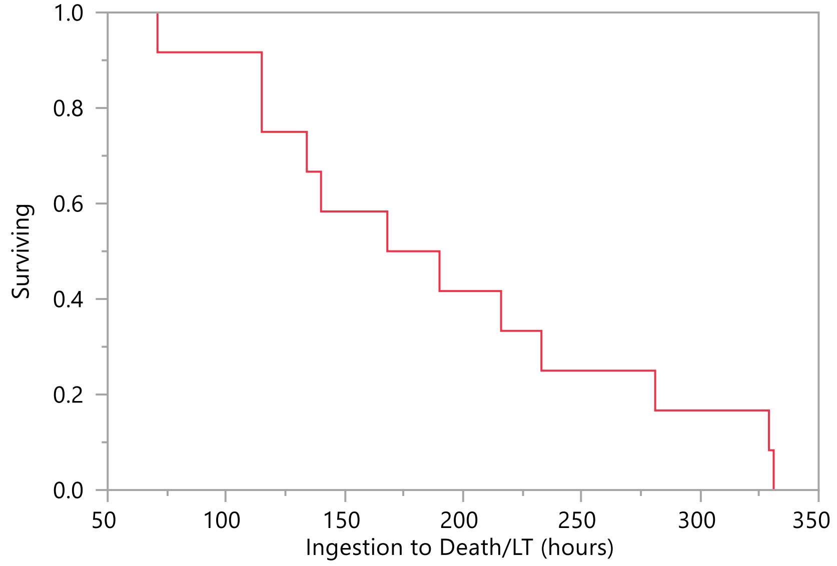

**Table S-1. Treatment Emergent Adverse Events (TEAEs)**
Safety Population (n=99)

| Body System | Organ System | Number | %[a] |
|---|---|---|---|
| Blood and lymphatic system disorders | | | |
| | Thrombocytopenia | 1 | 1.01% |
| Cardiac disorders | | | |
| | Supraventricular tachycardia | 1 | 1.01% |
| Gastrointestinal disorders | | | |
| | Bowel perforation | 1 | 1.01% |
| | Nausea | 1 | 1.01% |
| | Vomiting | 1 | 1.01% |
| General disorders and administration site conditions | | | |
| | Infusion site extravasation | 1 | 1.01% |
| | Pyrexia | 1 | 1.01% |
| Genitourinary disorders | | | |
| | Acute renal failure | 1 | 1.01% |
| Hepatobiliary disorders | | | |
| | Acute hepatic failure | 3 | 3.03% |
| Investigations | | | |
| | Osmolar gap abnormal | 1 | 1.01% |
| Nervous system disorders | | | |
| | Headache | 1 | 1.01% |
| | Hepatic encephalopathy | 1 | 1.01% |
| Psychiatric disorders | | | |
| | Mental status changes | 1 | 1.01% |
| Skin and subcutaneous tissue disorders | | | |
| | Dermatitis contact | 1 | 1.01% |
| Surgical and medical procedures | | | |
| | Hospitalization | 1 | 1.01% |
| Vascular disorders | | | |
| | Flushing | 6 | 6.06% |
| | Hypertension | 1 | 1.01% |

[a] *% of 99 patients with adverse events after intravenous silibinin initiation*

**Figure S-3. Severity, Relatedness and Action for TEAEs**

Safety Population (n=99)

Investigators reported 24 treatment emergent adverse events (TEAEs) involving 14 patients. Nine of these were judged serious adverse events (SAEs), of which 8 were judged not related and 1 possibly related.

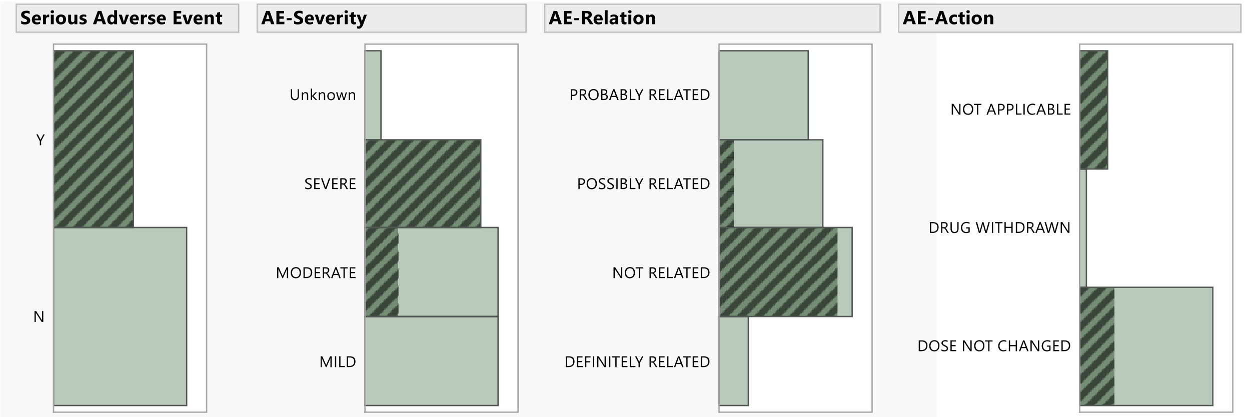

**Serious Adverse Event – Frequencies**

| Level | Count | Prob |
|---|---|---|
| N | 15 | 0.62500 |
| Y | 9 | 0.37500 |
| Total | 24 | 1.00000 |

N Missing 0
2 Levels

**AE-Severity – Frequencies**

| Level | Count | Prob |
|---|---|---|
| MILD | 8 | 0.33333 |
| MODERATE | 8 | 0.33333 |
| SEVERE | 7 | 0.29167 |
| Unknown | 1 | 0.04167 |
| Total | 24 | 1.00000 |

N Missing 0
4 Levels

**AE-Relation – Frequencies**

| Level | Count | Prob |
|---|---|---|
| DEFINITELY RELATED | 2 | 0.08333 |
| NOT RELATED | 9 | 0.37500 |
| POSSIBLY RELATED | 7 | 0.29167 |
| PROBABLY RELATED | 6 | 0.25000 |
| Total | 24 | 1.00000 |

N Missing 0
4 Levels

**AE-Action – Frequencies**

| Level | Count | Prob |
|---|---|---|
| DOSE NOT CHANGED | 19 | 0.79167 |
| DRUG WITHDRAWN | 1 | 0.04167 |
| NOT APPLICABLE | 4 | 0.16667 |
| Total | 24 | 1.00000 |

N Missing 0
3 Levels

### Figure S-4 Multivariate Model for Time to INR Recovery[a]

TFR + Recovery occurring after SIL started (n=64)

#### Effect Summary

| Source | Logworth | PValue |
|---|---|---|
| Ingestion to Symptoms (hours) | 1.810 | 0.01547 |
| Octreotide used | 1.486 | 0.03268 |

#### Regression Plot

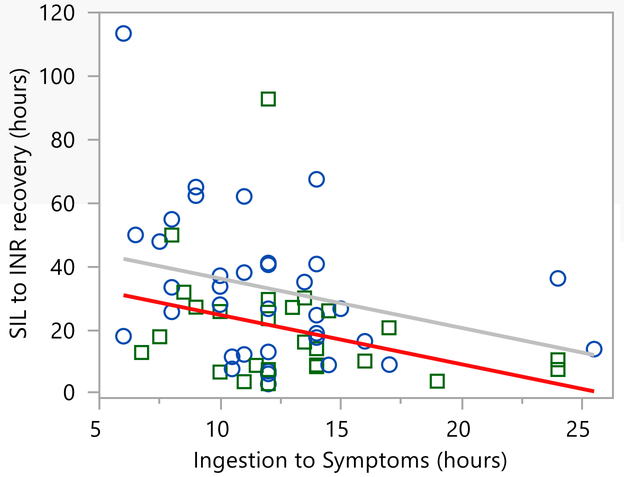

#### Summary of Fit

| | |
|---|---|
| RSquare | 0.170015 |
| RSquare Adj | 0.142803 |
| Root Mean Square Error | 20.57857 |
| Mean of Response | 27.4224 |
| Observations (or Sum Wgts) | 64 |

#### Ingestion to Symptoms (hours)

##### Leverage Plot

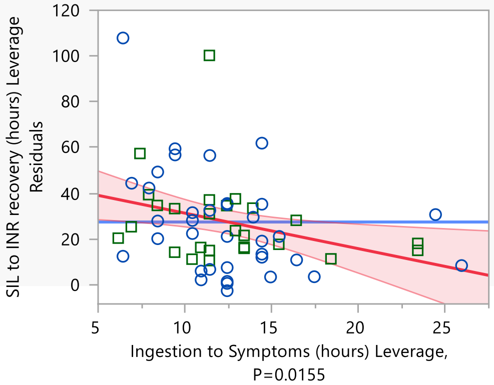

#### Octreotide used

##### Leverage Plot

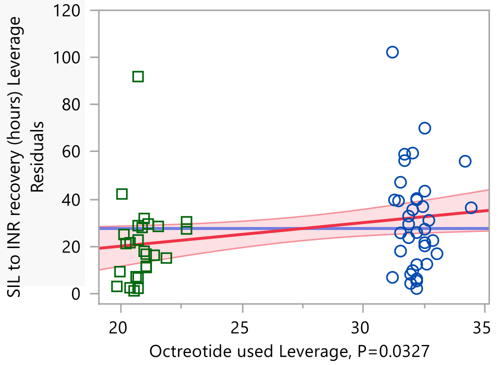

##### Least Squares Means Table

| Level | Least Sq Mean | Std Error | Mean |
|---|---|---|---|
| 0 | 32.2615 | 3.394 | 32.933 |
| 1 | 20.7910 | 3.978 | 19.870 |

#### Parameter Estimates

| Term | Estimate | Std Error | t Ratio | Prob>\|t\| |
|---|---|---|---|---|
| Intercept | 51.661315 | 8.244121 | 6.27 | <.0001 * |
| Ingestion to Symptoms (hours) | -1.555352 | 0.624366 | -2.49 | 0.0155 * |
| Octreotide used[1-0] | -11.47056 | 5.247697 | -2.19 | 0.0327 * |

[a] Same model obtained for forward, and backward stepwise logistic regression. Other predictors considered: Ingestion to first lab, Total SIL dose (mg/kg), Study maturity, Age, NAC used, Charcoal used, Bile drainage, and Sex.

**Table S-2. Hospital Length of Stay Relation to Patient and Treatment Measures**

No individual measure or multivariate model predicted hospital length of stay

| Binary Measure | -------- LOS Mean ± SEM [95% CI] (number) ------- |  | Difference [95% CI] |
|---|---|---|---|
| | Used/Present (1) | Not used/Absent (0) | (Used - Not used) |
| NAC used | 6.68±0.307 [6.07, 7.29] (66) | 5.79±0.572 [4.65, 6.93] (19) | 0.893 [-0.398, 2.18] |
| Charcoal used | 6.74±0.357 [6.03, 7.46] (49) | 6.13±0.417 [5.3, 6.96] (36) | 0.619 [-0.474, 1.71] |
| Bile drainage | 5.50±1.256 [3.00, 8.00] (4) | 6.53±0.279 [5.98, 7.09] (81) | -1.03 [-3.59, 1.53] |
| Sex (1=M, 0=F) | 6.64±0.345 [5.96, 7.33] (53) | 6.22±0.444 [5.34, 7.1] (32) | 0.424 [-0.7, 1.54] |
| Octreotide used | 6.37±0.426 [5.53, 7.22] (35) | 6.56±0.356 [5.85, 7.27] (50) | -0.187 [-1.29, 0.917] |

*p-value NS for all 5 binary measures [95% CI difference includes 0]*

| Continuous Measure | LOS slope [95% CI] |
|---|---|
| Age (years) | -0.0086 [-0.0349, 0.0178] |
| Ingestion to Treatment (hours) | -0.0083 [-0.0352, 0.0187] |
| Study maturity (years from first patient) | -0.0333 [-0.2256, 0.159] |

*p-value NS for all 3 continuous measures [95% CI includes 0]*

### Figure S-5 Maximum Laboratory Values versus TFR

Bivariate Logistic Fits of TFR Outcomes to Max Lab Values, Safety Population (n=99)

##### Logistic Fit of Outcome-2 By Max(Lactate)

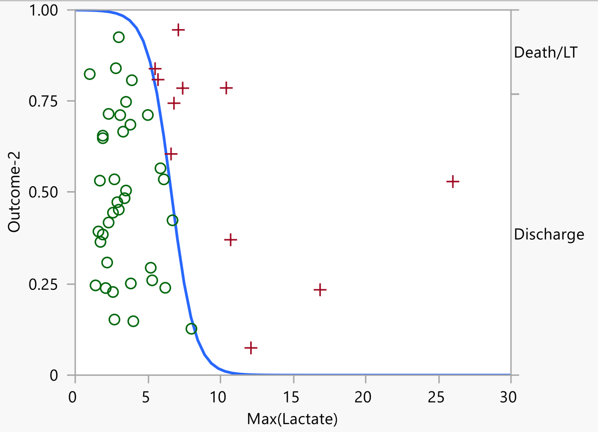

**Whole Model Test**

| Model | -LogLikelihood | DF | ChiSquare | Prob>ChiSq |
|---|---|---|---|---|
| Difference | 16.106031 | 1 | 32.21206 | <.0001 * |
| Full | 9.730807 | | | |
| Reduced | 25.836838 | | | |

| | |
|---|---|
| RSquare (U) | 0.6234 |
| AICc | 23.7283 |
| BIC | 27.204 |
| Observations (or Sum Wgts) | 48 |

**Parameter Estimates**

| Term | Estimate | Std Error | ChiSquare | Prob>ChiSq |
|---|---|---|---|---|
| Intercept | 8.15359098 | 2.8316666 | 8.29 | 0.0040 * |
| Max(Lactate) | -1.2327471 | 0.4591955 | 7.21 | 0.0073 * |

For log odds of Discharge/Death/LT

##### Logistic Fit of Outcome-2 By Max(INR)

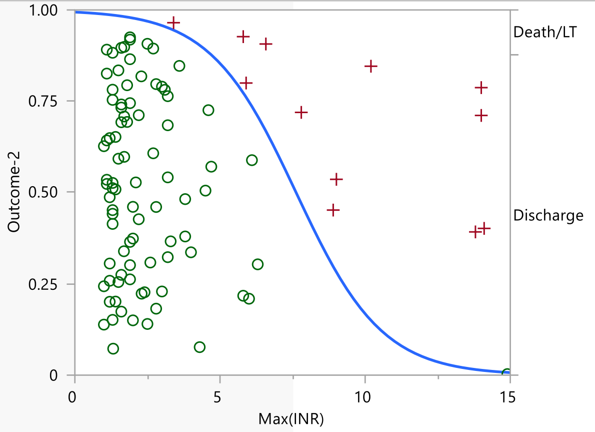

**Whole Model Test**

| Model | -LogLikelihood | DF | ChiSquare | Prob>ChiSq |
|---|---|---|---|---|
| Difference | 19.570964 | 1 | 39.14193 | <.0001 * |
| Full | 16.993015 | | | |
| Reduced | 36.563979 | | | |

| | |
|---|---|
| RSquare (U) | 0.5353 |
| AICc | 38.111 |
| BIC | 43.1763 |
| Observations (or Sum Wgts) | 99 |

**Parameter Estimates**

| Term | Estimate | Std Error | ChiSquare | Prob>ChiSq |
|---|---|---|---|---|
| Intercept | 5.12927022 | 0.9863282 | 27.04 | <.0001 * |
| Max(INR) | -0.6730061 | 0.1741929 | 14.93 | 0.0001 * |

For log odds of Discharge/Death/LT

Summary of Logistic Fits of TFR to Max Lab Values, Safety Population (n=99)
Sorted by RSquare

| Lab Max | RSquare (U) | Number of Patients | Chi-Square | Prob> ChiSq | Estimate | Std Error of Estimate |
|---|---|---|---|---|---|---|
| Lactate | 0.6234 | 48 | 7.207 | 0.00726 | -1.23275 | 0.45920 |
| INR | 0.5353 | 99 | 14.927 | 0.00011 | -0.67301 | 0.17419 |
| Total Bili | 0.1859 | 97 | 11.954 | 0.00055 | -0.18779 | 0.05432 |
| AST | 0.0701 | 99 | 5.286 | 0.02149 | -0.00014 | 0.00006 |
| Cr | 0.0580 | 96 | 4.376 | 0.03644 | -0.37004 | 0.17689 |
| BUN | 0.0334 | 96 | 2.518 | 0.11256 | -0.03102 | 0.01955 |
| ALT | 0.0287 | 99 | 2.226 | 0.13566 | -0.00010 | 0.00006 |
| Hct | 0.0137 | 95 | 0.994 | 0.31886 | -0.05093 | 0.05110 |
| Hgb | 0.0106 | 95 | 0.778 | 0.37768 | -0.12338 | 0.13985 |

#### Patient Narratives, Exclusion and Poor Outcome Patients

Patient 1:
30-40 y/o HF transferred from a community hospital ER to a transplant center after a 9-hour latency from ingestion to onset of symptoms. Presented (first labs) **53 hours** post-ingestion with Hgb 17.2, INR 2.2 (peak 3.4), **Lactate 5 (never corrected)**. Following admission to a tertiary transplant center **IV hydration was restricted** with **persistent lactate elevation** and **oliguric AKI**. FHF and multi-system failure developed quickly thereafter. Died on hospital day 13.

Patient 2:
30-40 y/o HF transferred from a community hospital ER to a transplant center after a 9-hour symptom latency following ingestion. Presented **53 hours** post-ingestion with Hgb 8.9, INR 8.9 (peak), **Lactate 5.7 (never corrected)**. Upon admission to transplant center **IV hydration restricted** with **persistent lactate elevation** and **oliguric AKI**. Successful liver transplant on hospital day 12.

Patient 3:
30-40 y/o HM transferred from a community hospital ER to a transplant center. Nine-hour latency between ingestion and onset of symptoms. Presented **51 hours** post- ingestion with INR 4.6 (peak), **Lactate 6.1**. Following admission to transplant center, **IV hydration was restricted**. **Lactate remained uncorrected and developed oliguric AKI**. Recovery and unremarkable hospital course with resolution of coagulopathy and oliguric AKI after undergoing **ERCP nasobiliary drainage** at transplant center <18 hours after ER presentation.

Patient 21:
60-70 y/o WM; first lab results ~**46 hours** post-ingestion but **did not begin SIL until 123 hours post-ingestion**. Twelve-hour latency between ingestion and onset of symptoms. Presentation Hgb 18.7, INR 1.0 (peak 5.8), Cr 2.3. Subsequent Cr values 1.1-1.9. At 102 hours post- ingestion, **Lactate 5.5 (never corrected)**. Developed **oliguric AKI** 101 hours following admission. Successful liver transplant on hospital day 8.

Patient 24:
40-50 y/o Eastern European F presented **104 hours** post-ingestion after a 24-hour symptom latency with Hgb 11.4, Cr 0.5, INR 3.8 (peak 14.1) and **Lactate 8.2 (never corrected)**. Developed **oliguric AKI** 60 hours following presentation. **Began SIL 146 hours post-ingestion**. Died on hospital day 8.

Patient 30:
65-75 y/o Central Asian F presented **22 hours** post-ingestion after a 14-hour symptom latency with Hgb 13.9, INR 1.0 (peak 2.46), Cr 1.13. **Received one liter total of NS during the 6 hours she was under treatment in the ER.** Per PCC recommendations she was started on multidose reactivated charcoal, NAC, and Cimetidine by infusion. **Oliguria** was noted by nursing staff **<3 hours after reaching the medical floor** from the ER. 17 hours post-presentation she became hypotensive (SBP 60) and acidotic (ph 6.99) with **Lactate 4.5 (never corrected)**. Death occurred <60 hours after ER presentation.

Patient 34:
50-60 y/o East Asian F presented **42 hours** post-ingestion after a symptom latency of 12 hours with Hgb 19.2, INR 1.8 (peak 12.2), Cr 3.0, and **Lactate 16.86 (never corrected)**. **Presented with oliguric AKI and made no urine in response to IV fluid challenge**. Began SIL 54 hours post-ingestion. Death on hospital day 13.

Patient 37:
15-25 y/o WM presented **126 hours** post-ingestion after an estimated symptom latency of 24 hours with Hgb 17.1, Cr 0.74, INR 14, and **Lactate 3.6 (never corrected)**. **Began SIL 150 hours post-ingestion** but **received less than 24 hours of SIL**, undergoing successful liver transplant later on the same day SIL was initiated.

Patient 47:
55-65 y/o East Asian F presented **45 hours** post-ingestion after an estimated symptom latency of 10 hours with Hgb 16.9, Cr 2.1, INR 1.6 (peak 13.8), and **Lactate 10.2 (never corrected)**. SIL began 55 hours post-ingestion. Died on hospital day 4. **IV hydration interrupted** intermittently during the course of treatment and transfers.

Patient 76:
01–04 y/o HF presented **20 hours** post-ingestion after a symptom latency of 12 hours with Hgb 10.8, Cr 0.379. At 32 hours post-ingestion INR was 1.4 (peak 9.8) and **Lactate 9.2**. She received aggressive volume replacement with **correction of Lactate to 2.1 (42 hours) and 1.7 (44 hours)** before transfer to a pediatric transplant center. Lactate correction sustained at 2.2 (54 hours) after which **IV hydration was restricted.** At 66 hours, **Lactate 7.1 (never corrected)**. SIL initiated 38 hours post-ingestion. Subsequently, encephalopathic with elevated serum ammonia values. Underwent liver transplant on hospital day 6 with sequelae of **permanent brain injury**. Died 9 years later.

Patient 82:
50-60 y/o HM presented **52 hours** after ingestion following a symptom latency of 14 hours with Hgb 20.9, Cr 2.3, INR 5, and **Lactate 5.4 (never corrected)**. Received **minimal volume replacement and restricted IV hydration**. SIL infused for 11 hours. Transferred to University Hospital and underwent a successful liver transplant on 5th-day post-ingestion.

Patient 182:
75-85 y/o WF presented **31 hours** post-ingestion after a symptom latency of 10.5 hours with HGB 14.2, Cr 0.85, INR 1.1 (peak 1.6). **Unable to tolerate flushing during initial bolus infusion, so SIL discontinued**. Received sustained aggressive IV hydration and recovered without complication.

Patient 188:
80-90 y/o HF presented **14 hours** post-ingestion after a symptom latency of 11 hours with Hgb 13.4, Cr 1.2 (peak 5.6), INR 0.9 (peak 5.9). Received aggressive IV hydration in community hospital. **Maintenance IV fluids discontinued** upon arrival at transplant center. Soon thereafter developed **oliguric AKI**. Died on hospital day 10.

Patient 11: (**SAE: AKI-ATN**) (not excluded)
35-45 y/o WF with history of IDDM and HTN on Lisinopril had a 10-hour latency between ingestion and first symptoms. Presented **37 hours** post-ingestion with Lactate 2.7, Hgb 16.9, BUN 28, Cr 1.1, CO2 14, INR 1.1 (Peak 2.4). Developed **non-oliguric AKI-ATN** (positive biopsy) with peak Cr 8.83, decreasing to 6.68 on day of discharge with resolution several months later (no HD).
AKI-ATN developed as FHF was resolving.

Patient 71 (**SAE: AKI-ATN**) (not excluded)
50-60 y/o South Asian M presented ~ **54 hours** post-ingestion after latency 12+ hours with BUN 42, Cr 4.79, CO2 9 (8/0.93/25 at discharge ~ 54 hours later). Treated ~ 48 hours with SIL and recovered with normal BUN/Creat and recovering INR. **Readmitted 11 days later** with peripheral and pulmonary edema, **non-oliguric AKI**. Renal biopsy demonstrated **ATN** (Cr 8.69; peak 9.13). Underwent HD x 2. At the second hospital discharge 8 days later, Cr 3.95 and trending down.

Patient 79 (not excluded):
35-45 y/o HF presented **36 hours** post-ingestion after a symptom latency of <12 hours with Hgb 15.9, Cr 0.8, INR 1.8 (peak 7.8), and **Lactate 7.0 which never fully corrected** despite aggressive IV volume replacement (also had **persistent non-correcting sinus tachycardia**). Received ~36 hours of SIL infusion before transfer to a University Hospital and undergoing successful liver transplantation 2 days later.

**SUPPLEMENTARY APPENDIX — Table S4**

**Persistent Misconceptions in Amatoxin Poisoning: Evidence-Based Clarification**

| Misconception | Current Status | Evidence-Based Clarification | References |
|---|---|---|---|
| Phallotoxins cause the gastrointestinal syndrome | Still cited in textbooks and reviews | The GI syndrome is reproduced by parenteral α-amanitin alone in dogs; phallotoxins are not absorbed from the GI tract and do not contribute to clinical toxicity | Fiume 1973; Faulstich Fauser 1980 |
| 60% of amatoxin undergoes enterohepatic recycling | Widely quoted figure | No identifiable primary source in the literature; likely overstates EHC contribution when renal clearance is preserved (>80% cleared renally under intact perfusion) | No primary reference identified |
| N-acetylcysteine improves outcomes | Routinely recommended; considered standard of care | No toxicokinetic rationale (amatoxin eliminated intact, not metabolized); no benefit demonstrated in animal models; marginal benefit in one adult non-acetaminophen ALF trial that excluded amatoxin patients; transient INR elevation may confound prognostic assessment | This trial; Lee et al. 2009; Cochrane reviews |
| Activated charcoal is effective | Standard recommendation, | Amatoxin's polarity, water solubility, and rapid absorption | This trial; Juurlink 2016 |

| | | | |
|---|---|---|---|
| | including multi-dose regimens | make it a poor charcoal substrate; intestinal toxin burden low after absorption complete until meal-triggered EHC; aspiration and ileus risks | |
| Penicillin G improves outcomes | Widespread historical use; still recommended in some protocols | No outcome association demonstrated in two large multivariable analyses despite being the most frequently used intervention | Enjalbert et al. 2002; Poucheret et al. 2010 |
| Fluid restriction prevents cerebral edema in amatoxin ALF | Standard transplant-unit practice for other ALF etiologies | Contraindicated in amatoxin poisoning; compromises renal perfusion and the dominant elimination pathway; multiple poor outcomes followed transplant-unit hydration reduction | This trial; patient narratives in Supplementary Appendix |
| Oral silymarin can substitute for IV silibinin | Sometimes used when IV formulation unavailable | Oral silymarin is poorly absorbed; a recent controlled study failed to establish efficacy | Tangsuwanaruk et al. 2026 |
| Renal replacement therapy can substitute for native kidney function | Often initiated for ammonia clearance or "renal support" | Native kidneys provide rapid amatoxin clearance that no extracorporeal modality can match; RRT may precipitate oliguric AKI if accompanied by reduced IV hydration | Jaeger 2004. This trial |

| | | | |
|---|---|---|---|
| Hepatic uptake inhibitors with nephrotoxic properties are acceptable alternatives | Proposed as silibinin alternatives | Agents that impair renal perfusion and glomerular filtration may precipitate oliguric AKI, compromising the dominant elimination pathway | Pharmacologic principles; this trial framework |

This table addresses common misconceptions that persist in clinical practice guidelines, textbooks, and online resources despite limited or contradictory evidence. Many of these practices were established before the toxicokinetic framework presented in this trial was understood. The clarifications provided are based on primary literature review, animal studies, retrospective analyses, and findings from this prospective trial. Adoption of evidence-based management requires recognition that interventions lacking toxicokinetic rationale may displace elimination-critical priorities. GI, gastrointestinal; EHC, enterohepatic circulation; ALF, acute liver failure; INR, international normalized ratio; AKI, acute kidney injury; RRT, renal replacement therapy; IV, intravenous.
